# One-carbon metabolism-related compounds are associated with epigenetic aging biomarkers: Results from National Health and Nutrition Examination Survey (NHANES) 1999-2002

**DOI:** 10.1101/2025.01.06.25320074

**Authors:** Anne Bozack, Dennis Khodasevich, Jamaji C. Nwanaji-Enwerem, Nicole Gladish, Hanyang Shen, Saher Daredia, Mary Gamble, Belinda L. Needham, David H. Rehkopf, Andres Cardenas

**Affiliations:** Department of Epidemiology and Population Health, Stanford University, Palo Alto, CA, US; Department of Emergency Medicine, Center for Health Justice, and Center of Excellence in Environmental Toxicology, Perelman School of Medicine, University of Pennsylvania, Philadelphia, PA, US; Department of Epidemiology, School of Public Health, University of California, Berkeley, Berkeley, CA, US; Department of Environmental Health Sciences, Mailman School of Public Health, Columbia University, New York, NY, US; Department of Epidemiology, University of Michigan, Ann Arbor, Michigan, US; Department of Health Policy, Stanford University, Palo Alto, CA, USA; Department of Medicine - Primary Care and Population Health, Stanford University, Palo Alto, CA, US; Department of Pediatrics, Stanford University, Palo Alto, CA, USA; Department of Sociology, Stanford University, Palo Alto, CA, USA

**Keywords:** one-carbon metabolism, epigenetic aging, DNA methylation, folate, homocysteine, vitamin B12, National Health and Nutrition Examination Survey (NHANES)

## Abstract

**Background:** One-carbon metabolism (OCM), a biochemical pathway dependent on micronutrients including folate and vitamin B12, plays an essential role in aging-related physiological processes. DNA methylation-based aging biomarkers may be influenced by OCM.

**Objective:** This study investigated associations of OCM-related biomarkers with epigenetic aging biomarkers in the National Health and Nutrition Examination Survey (NHANES).

**Methods:** Blood DNA methylation was measured in adults aged ≥50 years in the 1999-2000 and 2001-2002 cycles of NHANES. The following epigenetic aging biomarkers were included: Horvath1, Horvath2, Hannum, PhenoAge, GrimAge2, DunedinPoAm, and DNA methylation telomere length (DNAmTL). We tested for associations of serum folate, red blood cell (RBC) folate, vitamin B12, homocysteine, and methylmalonic acid concentrations with epigenetic age deviation (EAD) among 2,346 participants with epigenetic and nutritional status biomarkers using survey weighted general linear regression models adjusting for sociodemographics, BMI, and behavioral factors.

**Results:** A doubling of serum folate concentration was associated with −0.82 years (95% confidence interval (CI) = −1.40, −0.23) lower GrimAge EAD, −0.13 SDs (−0.22, −0.03) lower DunedinPoAm, and 0.02 kb (0.00, 0.04) greater DNAmTL EAD. Associations were attenuated after adjusting for smoking status and alcohol intake, folate antagonists. Conversely, a doubling in homocysteine concentration was associated with 1.05 years (0.06, 2.04) greater PhenoAge EAD, 1.93 years (1.16, 2.71) greater GrimAge2 EAD, and 0.26 SDs (0.10, 0.41) greater DunedinPoAm. Associations with GrimAge2 EAD and DunedinPoAm were robust to alcohol and smoking adjustment.

**Conclusions:** In a nationally representative sample of U.S. adults, greater folate, a carbon donor, was associated with lower EAD, and greater homocysteine, an indicator of OCM deficiencies, was associated with greater EAD; however, some associations were influenced by smoking status. Future research should focus on high-risk populations. Randomized controlled trials with long-term follow-up are also needed to established causality and investigate the clinical relevance of changes in EAD.

## Background

One-carbon metabolism (OCM) is a biochemical pathway essential for supporting many physiological processes. OCM produces the universal methyl donor *S*-adenosylmethionine (SAM), a cofactor for methylation of DNA, proteins, amino acids, and other small molecules. OCM also includes the transsulfuration pathway, which generates the antioxidant glutathione, and provides one-carbon units for purine and thymidylate biosynthesis. OCM plays an essential role in aging-related processes by supplying methyl groups, producing building blocks for DNA synthesis and repair, and maintaining redox balance in cells.^1^ Deficiencies or imbalances in OCM increase the risk of aging-related physiological changes and diseases^1–3^ and may influence aging-related biomarkers.

OCM is dependent on diet-derived micronutrients including folate (vitamin B9) and cobalamin (vitamin B12)^4^ (**Figure 1**; adapted from^5^). Naturally occurring folates are found in foods such as vegetables, fruits, legumes, and nuts. Folic acid is a synthetic, stable form of folate used in supplements and fortified foods. Folate facilitates the recruitment of methyl groups to OCM, which are subsequently used to remethylate homocysteine (Hcy) to methionine, the precursor of SAM, by methionine synthase using B12 as a cofactor. Upon donation of a methyl group, SAM is converted to S-adenosylhomocysteine (SAH), a product inhibitor of most methylation reactions, which can be hydrolyzed to Hcy and remethylated to methionine given adequate folate and B12. B12 is also involved in the metabolism of methylmalonic acid (MMA), a byproduct of propionate metabolism.^6^ High Hcy concentration is an indicator of folate or B12 deficiency, and high MMA concentration is an indicator of B12 deficiency.^7,8^

**Figure 1:**
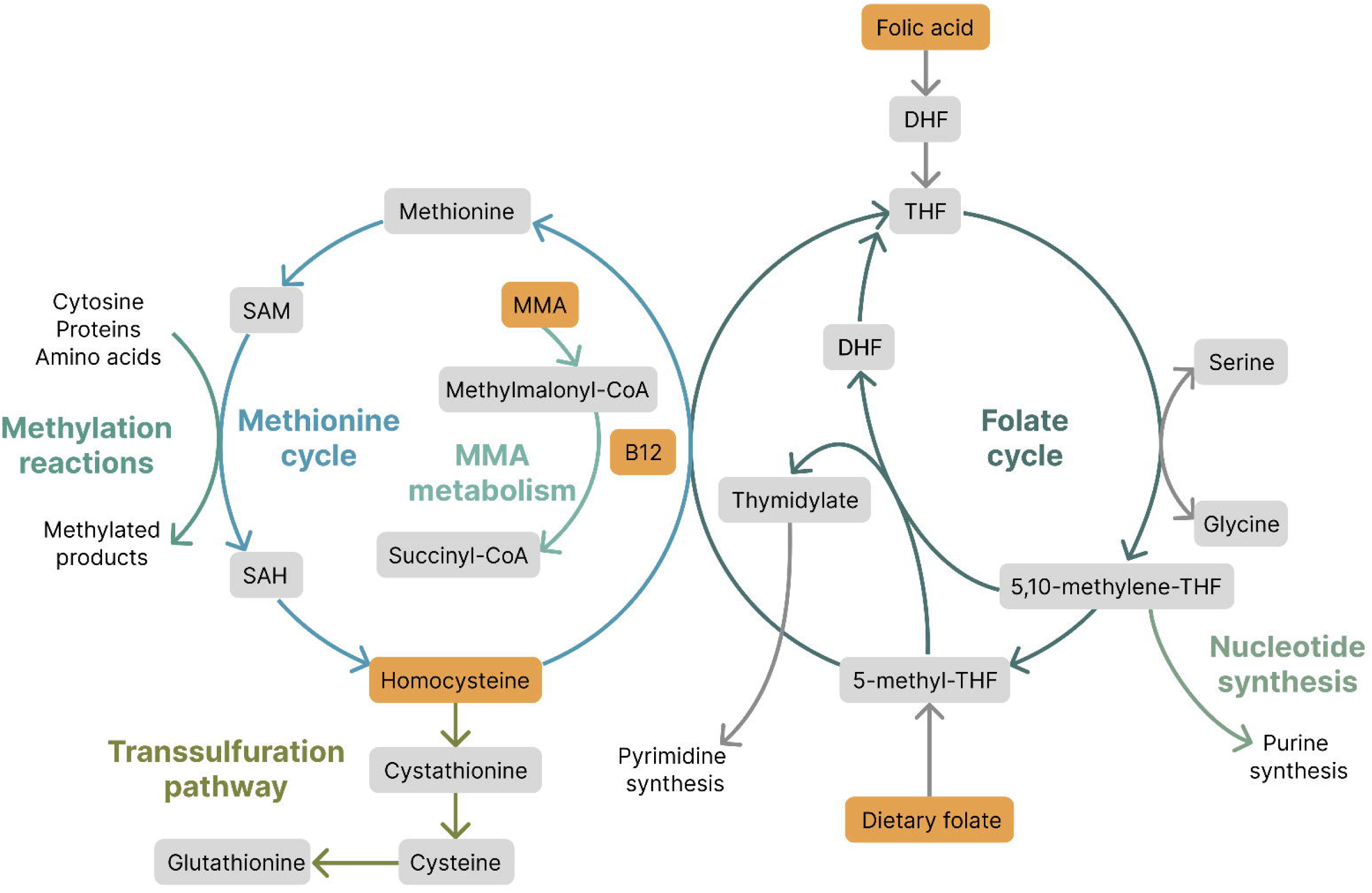
One-carbon metabolism. Compounds included in this study are shown in orange.

The prevalence of folate and B12 deficiencies increases with age due to changes in diet, intestinal absorption, and use of antagonistic therapeutic drugs.^9,10^ Low folate levels, which can have mutagenic effects due to reduced thymidylate biosynthesis and increased uracil misincorporation,^11^ have been associated with age-related conditions, including dementia,^12^ cognitive function,^13^ and cancers.^14,15^ Folate and B12 deficiencies also contribute to disease risk by increasing Hcy concentration. Hcy can induce oxidative stress, inflammation, and impaired endothelial function.^16^ Elevated Hcy has been associated with increased risk of cardiovascular disease (CVD) incidence and mortality,^17^ cognitive impairment and decline,^18^ particularly in groups with existing neurological disorders,^19–21^ and all-cause mortality,^17^ although associations may vary by population due to differences in diet, age, and genetic factors.^22^

In recent years, the study of aging and the healthspan has advanced with the development of epigenetic aging biomarkers, also known as epigenetic clocks. These biomarkers leverage DNA methylation levels at specific cytosine-guanine dinucleotides (CpG sites) that change with age. This phenomenon has been leveraged to train epigenetic clocks as linear combinations of DNA methylation levels at age-related CpGs. Epigenetic clocks can be broadly classified as predictors of chronological age (*i.e.*, first-generation clocks),^23,24^ aging-related phenotypes and mortality risk (*i.e.*, second-generation clocks),^25–27^ and rate-of-aging (*i.e.*, third generation clocks).^28^ Epigenetic age deviation (EAD), or the difference between epigenetic and chronological age, is strongly associated with cognitive impairment, dementia, BMI, cancer, CVD, other morbidities, and all-cause mortality.^29,30^ Second generation clocks have proven to be particularly strong predictors of cancer, CVD, comorbidity, and time to death^25,27^ — aging-related outcomes that may have etiologies related to OCM deficiencies.

Testing the relationship between markers of OCM and EAD can advance our understanding of how nutritional factors and OCM function are related to complex aging- and disease-related processes and can inform the application of epigenic aging biomarkers in research and clinical settings. In this study, we aimed to investigate associations of OCM-related micronutrients and biomarkers with EAD among participants in the National Health and Nutrition Examination Survey (NHANES), a nationally representative sample of adults in the United States (US). Our study focused on concentrations of the B vitamins folate and B12, Hcy, and MMA and seven epigenetic aging biomarkers (Horvath1,^23^ Horvath2,^24^ Hannum,^31^ PhenoAge,^25^ GrimAge2,^26^ DunedinPoAm,^28^ and DNA methylation telomere length (DNAmTL)^32^). We hypothesized that promoters of OCM (*i.e.*, folate and B12) would be associated with lower EAD, whereas indicators of OCM deficiencies (*i.e.*, Hcy and MMA) would be associated with increased EAD.

## Methods

### Study population

We leveraged data from the 1999-2000 and 2001-2002 cycles of NHANES. NHANES is an ongoing program conducted by the National Center for Health Statistics (NCHS) that includes interviews, physical exams, and laboratory measurements and is designed to evaluate and monitor the health and nutrition among noninstitutionalized adults and children in the US.^33^ In the 1999-2000 and 2001-2002 cycles of NHANES, DNA methylation was measured in blood samples of a subset of adults aged 50 years or older. To protect participant confidentiality, participants ≥ 85 years were top coded as 85 years old. Consequently, all participants assigned an age of 85 years were excluded from our analyses as we were unable to determine their true chronological age. Our study included 2,346 adults with available DNA methylation data and nutritional biomarkers after excluding sex-mismatches and participants 85 years and older (**Supplemental Figure 1**). All participants provided written informed consent, and the study protocols were approved by the NCHS Research Ethics Review Board.^34^

### DNA methylation

A detailed description of DNA methylation laboratory methods and data processing can be found on the NHANES website.^35^ In the 1999-2000 and 2001-2002 cycles of NHANES, DNA methylation was measured in blood samples of 2,532 adults. Samples were selected for DNA methylation measurement among participants aged 50 years or older with available biospecimens and included a random sample of approximately half of eligible non-Hispanic White participants and all eligible participants from other ethnic and racial groups (*i.e.*, non-Hispanic Black, Mexican American, other Hispanic, and other race).^35^ DNA was extracted from whole blood and stored at −80°C until DNA methylation measurement. DNA underwent bisulfite conversion and DNA methylation was measured using the Illumina Infinium MethylationEPIC BeadChip v1.0 (Illumina, San Diego, CA, US) following the manufacturer’s recommendations. The MethylationEPIC BeadChip interrogates over 850,000 CpG dinucleotides.^36^ Raw DNA methylation data were imported into R for preprocessing and quality control (QC). The preprocessing and QC pipeline included color correction, background subtraction, and removal of outlier samples based on median intensity values.

### Epigenetic age and biomarkers

Epigenetic age estimates and aging biomarkers were provided by NHANES.^35^ Prior to biomarker calculation, DNA methylation data were normalized using the beta mixture quantile (BMIQ) method, which adjusts for probe type bias.^37^ For the calculation of the Horvath1, Horvath2, PhenoAge, and GrimAge2 clocks, a modified BMIQ method was applied using a gold standard based on the largest training dataset in the Horvath1 clock.^23^ The epigenetic age estimates or scores were calculated for each participant using the respective published coefficients and intercepts. We focused on widely studied and established epigenetic aging biomarkers including three first-generation clocks (*i.e.*, trained to estimate chronological age): Horvath1,^23^ Horvath2,^24^ and Hannum;^31^ two second-generation clocks (*i.e.*, trained to predict aging-related phenotypes, mortality, and morbidity risk): PhenoAge^25^ and GrimAge2;^26^ a third-generation clock (*i.e.*, trained to measure the rate of change of epigenetic age): Dunedin Pace-of-Aging (DunedinPoAm);^28^ and telomere length (DNAmTL).^32^ GrimAge2 is an updated version of the GrimAge biomarker of mortality risk, which includes training on additional DNA methylation-based estimates of plasma proteins (high sensitivity C-reactive protein (CRP) and hemoglobin A1C). Due to the high correlation between GrimAge and GrimAge2 in NHANES (*r* = 0.99), we chose to only analyze GrimAge2. In addition to GrimAge2 calculations, available NHANES data also includes DNA methylation-based estimates of plasma proteins used in the calculation of this clock.

### Nutritional biochemistries

Methods for nutritional biochemistries are described on the NHANES website with the respective data releases.^38,39^ Briefly, serum folate and B12 concentrations were measured using the Quantaphase II Folate/vitamin B12 radioassay kit (Bio-Rad Laboratories, Hercules, CA, US).^38^ RBC folate was measured by diluting samples in ascorbic acid in water and incubating or freezing to hemolyze RBCs. Samples were diluted again in protein diluent and RBC folate was measured using methods analogous to serum folate. Plasma Hcy was measured using the Abbott Homocysteine assay, a fluorescence polarization immunoassay, and the Abbott IMX analyzer (Abbott Diagnostics, Chicago, IL, US). MMA was extracted from plasma or serum with a strong anion exchange resin, and concentrations were measured by gas chromatography and a mass selective detector.

### Covariates

Demographic, socioeconomic, anthropometric measures, and self-reports of health-related behaviors were collected as part of the NHANES questionnaire and physical examination. As defined by NHANES, chronological age at the time of the screening interview was calculated from reported date of birth. In the case of missing date of birth, reported age in years was used to impute date of birth. Self-reported race and ethnicity were classified as Non-Hispanic White, Mexican American, other Hispanic, Non-Hispanic Black, or other race including Multiracial. Smoking status was based on the Smoking and Tobacco Use Questionnaire Section and classified as never (not having smoked at least 100 cigarettes in life), former (having smoked at least 100 cigarettes in life but not currently smoking), or current (having smoked at least 100 cigarettes in life and currently smoking every day or some days). Alcohol intake in average number of drinks per day in the last 12 months was calculated from the Alcohol Use Questionnaire Section. Occupational status was obtained from the Occupation Questionnaire Section and classified as white-collar and professional work, white-collar and semi-routine work, blue-collar and high-skill work, blue-collar and semi-routine work, or no work as described.^40^ Education level was classified as less than high school, high school diploma (including GED), or more than high school. The poverty to family income ratio (PIR) was based on the Department of Health and Human Services’ (HHS) poverty guidelines and calculated as the family income divided by the poverty guidelines for family size and year. Body mass index (BMI) in kg/m^2^ was calculated from height and weight measurements collected by trained technicians during examinations.

Data on selected protein concentrations measured in blood were also available in NHANES. Serum β-2 microglobulin (B2M) concentrations were measured with a B2M immunoassay and serum cystatin C concentrations were measured with a Cystatin C immunoassay on an automated multi-channel analyzer (Siemens Healthcare Diagnostics, Tarrytown, NY, US).^41^ CRP concentrations were measured using latex-enhanced nephelometry.^42,43^ Glycohemoglobin (hemoglobin A1C) concentrations in plasma were measured using an HPLC-based automated glycohemoglobin analyzer (Primus, Kansas City, MO, US).^44,45^

### Statistical analyses

Among 2,532 participants with epigenetic age estimates, we excluded those with a recorded age of ≥85 years (*n* = 130) or with a mismatch between recorded sex and a DNA methylation-based sex estimate (*n* = 56) (**Supplementary Figure 1**). We calculated epigenetic age deviation (EAD), also known as epigenetic age acceleration,^46^ as the residuals of regressing chronological age in years on epigenetic age. DunedinPoAm was analyzed centered and scaled. Blood cell type proportions (CB8+ T cells, CD4+ T cells, neutrophils, monocytes, B cells, and natural killer cells) were estimated using regression calibration^47^ based on IDOL probe selection.^48,49^

Descriptive statistics before survey weighting were calculated as means and standard deviations (SDs) for continuous variables and frequencies and proportions for categorical variables. We evaluated performance of the epigenetic aging biomarkers by calculating correlations and median absolute errors (MAEs) between chronological age and estimated epigenetic age. Survey weights were provided with the NHANES epigenetic biomarkers dataset.^35^ The survey design was specified using the *Survey* R package for statistical analyses.^50,51^ To preserve the study design, the survey design was specified prior to dropping participants ≥ 85 years old and sex mismatches. Correlations between OCM-related micronutrient concentrations (serum folate, RBC folate, B12, Hcy, and MMA) were calculated using the *svycor* function using the bootstrap procedure in the *jtools* R package.^52^

We evaluated associations of OCM-related micronutrients and biomarkers with epigenetic aging biomarkers (EAD residuals or DunedinPoAm) using survey-design weighted generalized linear regression models. Analyses were conducted using the *svyglm* function in the *Survey* R package.^50,51^ The concentrations of serum folate, RBC folate, B12, Hcy, and methylmalonic acid were log_2_ transformed to reduce the influence of outliers and meet model assumptions. Primary models were adjusted for potential confounders and precision variables identified *a priori*: chronological age, chronological age^2^, sex, race and ethnicity, smoking status, education level, occupation, and PIR.

Smoking and alcohol intake may be associated with concentrations of OCM-related compounds due to lifestyle and dietary factors. In addition, smoking^53^ and alcohol^54^ may act as folate antagonists by affecting the absorption, metabolism, and maintenance of circulating levels. In our NHANES study sample, we found that current smoking status compared to never smoking was negatively associated with the concentrations of serum folate, RBC folate, and MMA and positively associated with the concentration of Hcy (*p* < 0.05; **Supplemental Tables 1 and 2**), consistent with previous studies.^55,56,56^ Alcohol intake was negatively associated with B12 concentration and positively associated with Hcy concentration (*p* < 0.05). In addition, former and current smokers reported greater alcohol intake (drinks/day, never smokers = 0.2; former smokers = 0.4; current smokers = 0.8; p < 0.001) (**Supplemental Table 1**). Therefore, we also conducted analyses adjusting for smoking status (former or current smoking vs. never) and alcohol intake to evaluate if associations of OCM-related compounds and epigenetic aging biomarkers were independent of these exposures. When adjusting for smoking or alcohol alone, we saw the largest change in effect sizes with inclusion of smoking in the model (data not shown), and, consequently, we also tested for interactions between the concentrations of OCM-related compounds and smoking status by conducting stratified analyses and including an interaction term in unstratified analyses.

We further added log_2_-transformed cystatin C concentration, an indicator of renal function, as a covariate to the models. Elevated homocysteine can impair renal function, and, in turn, impaired renal function increases the risk of cardiovascular disease.^57^ In addition, poor renal function has been associated with increased serum folate and RBC folate concentration in a previous study in NHANES.^58^

Considering conflicting evidence of the relationship between excess folate and B12 and adverse health outcomes, including cancer progression and cognition,^59–63^ we tested for non-linear associations using fully adjusted models with folate tertiles (serum folate: < 11.2, ≥ 11.2 and < 17.6, and ≥ 17.6 ng/mL; RBC folate: < 250, ≥ 250 and < 352, and ≥ 352 ng/mL RBC) and B12 tertiles (< 398, ≥ 398 and < 583, and ≥ 583 pg/mL). We also conducted analyses with the dichotomous variable hyperhomocysteinemia (Hcy concentration > 15 umol/L)^64^ vs. not.

To evaluate if individual GrimAge2 components were contributing to the associations with GrimAge2 EAD, we conducted adjusted analyses of serum folate, RBC folate, and homocysteine concentrations using GrimAge2 components (plasma protein surrogates and smoking pack years) as the outcomes. We conducted complementary analyses for proteins with laboratory measures, if available (B2M, cystatin C, CRP, and hemoglobin A1C).

We performed sensitivity analyses adjusting for cell type proportions to distinguish between intrinsic aging (*i.e.*, intracellular) and extrinsic aging (*i.e.*, reflecting age-related changes in cell type proportions). Our primary analyses were conducted using complete cases. We also conducted sensitivity analyses following imputation of missing covariate data. Data were imputed with multiple imputation by chained equations using the *MICE* R package^65^ and 5 iterations. Following imputation, we reanalyzed associations of OCM-related micronutrients and compounds with aging biomarkers using fully adjusted weighted generalized linear regression models and pooled estimates from each imputed dataset with the *pool* function.

We used 95% confidence intervals (95% CIs) to evaluate precision of associations and *p* < 0.05 to test for statistical significance. All analyses were conducted in R version 4.4.1.^66^ We used the STROBE-nut checklist when writing our report.^67^

## Results

Epigenetic aging biomarker and nutritional biomarker data were available for 2,346 participants. Participant demographic characteristics and concentrations of OCM-related nutrients and compounds, prior to survey weighting, are shown in **Table 1**. Participants had a mean (SD) chronological age of 65.1 (9.3) years, and approximately half (51.2%) of participants were male. Nearly all participants were folate replete; 5 participants had serum folate concentrations less than or equal to the reference value of 3 ng/mL.^68^ Twenty-one (0.9%) participants were classified as B12 deficient (concentration <150 pg/mL) and 329 (14.0%) were classified as B12 insufficient (concentration < 300 pg/mL).^8^ Hyperhomocysteinemia was more common, with 192 (8.2%) participants having Hcy concentration > 15 umol/L.^64^

**Table 1:** Participant characteristics (*N* = 2,346).

|  | n (%) or mean (SD) |
| --- | --- |
| Male, n (%) | 1,202 (51.2%) |
| Age at screening, mean (SD) | 65.1 (9.3) |
| Race and ethnicity, n (%) |  |
| Mexican American | 681 (29.0%) |
| Other Hispanic | 151 (6.4%) |
| Non-Hispanic White | 922 (39.3%) |
| Non-Hispanic Black | 511 (21.8%) |
| Other race and Multiracial | 81 (3.5%) |
| BMI (kg/m<sup>2</sup>), mean (SD) | 28.8 (5.8) |
| Missing, n (%) | 84 (3.6%) |
| Smoking status, n (%) |  |
| Current | 374 (15.9%) |
| Former | 903 (38.5%) |
| Never | 1,063 (45.3%) |
| Missing | 6 (0.3%) |
| Alcohol intake, drinks/day, mean (SD) | 0.4 (1.0) |
| Missing, n (%) | 111 (5.0%) |
| Serum folate (ng/mL), mean (SD) | 16.5 (12.2) |
| Missing, n (%) | 1 (<0.01%) |
| Folate deficiency <sup>a</sup> | 5 (<0.01%) |
| Red blood cell folate (ng/mL RBC), mean (SD) | 328 (159) |
| Missing, n (%) | 5 (0.2%) |
| Vitamin B12 (pg/mL), mean (SD) | 734 (4,840) |
| Missing, n (%) | 1 (0.0%) |
| Vitamin B12 insufficiency, n (%) <sup>b</sup> | 329 (14.0%) |
| Vitamin B12 deficiency, n (%) <sup>c</sup> | 21 (0.9%) |
| Homocysteine (umol/L), mean (SD) | 10.1 (5.87) |
| Missing, n (%) | 3 (0.1%) |
| Hyperhomocysteinemia <sup>d</sup> | 192 (8.2%) |
| Methylmalonic acid (umol/L), mean (SD) | 0.2 (0.8) |
| Missing, n (%) | 7 (0.3%) |
| a. Serum folate ≤ 3 ng/mL. b. B12 < 300 pg/mL. c. < 150 pg/mL. d. > 15 umol/L. |  |

Participant characteristics stratified by smoking status are shown in **Supplemental Table 1**. Compared to never smokers, former smokers were more likely to be male and had greater alcohol intake, lower median B12 concentration, and greater median Hcy concentration; current smokers were more likely to be male and younger, and had lower BMI, greater alcohol intake, lower median concentration of serum folate and RBC folate, and higher median concentration of Hcy.

All first- and second-generation clocks were strongly correlated with chronological age, with Pearson correlation coefficients ranging from *r* = 0.76 for PhenoAge to *r* = 0.87 for Horvath2 (*p* < 0.001) (**Supplemental Figure 2**). The Horvath2 clock had the lowest MAE compared to chronological age (2.71 years) whereas PhenoAge had the largest MAE (10.33 years). As expected, DunedinPoAm was very weakly correlated with chronological age (*r* = 0.04; *p* = 0.049) and DNAmTL was negatively correlated with chronological age (*r* = −0.58; *p* < 0.001).

After survey weighting, there were moderate correlations of serum folate with RBC folate and Hcy with MMA (*r* = 0.51 and 0.50, respectively; *p* < 0.001) (**Supplemental Figure 3**). Serum folate was significantly but weakly associated with B12 (*r* = 0.04; *p* = 0.034), Hcy (*r* = −0.07; *p* = 0.002), and MMA (*r* = 0.05; *p* = 0.014). There were also weak correlations of RBC folate with Hcy (*r* = −0.09; *p* < 0.001) and B12 with Hcy (*r* = −0.05; *p* <0.001) and MMA (*r* = −0.02; *p* < 0.001).

### Associations of folate and B12 with epigenetic aging biomarkers

Figure 2 and **Supplementary Table 3** show results of our analyses of promoters of OCM (*i.e.*, serum and RBC folate and B12). Our primary analyses used weighted generalized linear regression models adjusted for chronological age, chronological age^2^, sex, race and ethnicity, BMI, education level, occupation, and PIR. We additionally adjusted for self-reported smoking status, alcohol intake, and cystatin C, an indicator of renal function.

**Figure 2:**
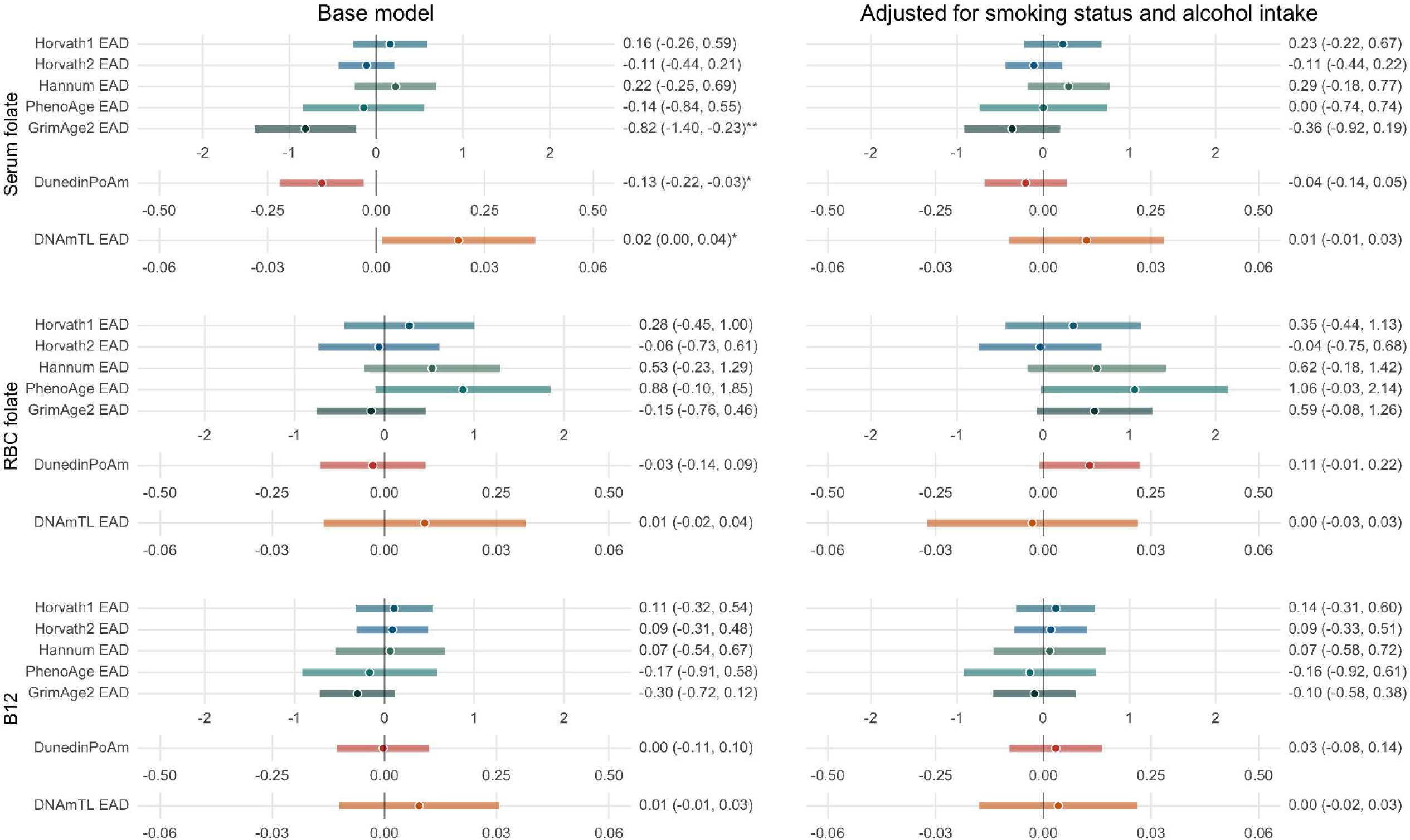
Associations of promoters of one-carbon metabolism (OCM) with epigenetic aging biomarkers. Effect estimates (95% confidence intervals (CIs)) and *p*-values are shown for a doubling in concentration of each compound. Results are from weighted generalized linear regression models adjusted for age, age^2^, sex, race and ethnicity, BMI, education level, occupation, and poverty to income ratio. Effect estimates for Horvath1, Horvath2, Hannum, PhenoAge, and GrimAge2 EAD are in years; effect estimates for DunedinPoAm are in standard deviations; and effect estimates for DNAmTL EAD are in kilobases. RBC = red blood cell; EAD = epigenetic age deviation. * *p* < 0.05; ** *p* < 0.01; *** *p* < 0.001.

Each doubling of serum folate concentration was associated with −0.82 years (95% CI = −1.40, −0.23; *p* = 0.010) lower GrimAge EAD, −0.13 SDs (95% CI = −0.22, −0.03; *p* = 0.015) lower DunedinPoAm, and 0.02 kb (95% CI = 0.00, 0.04; *p* = 0.037) greater DNAmTL EAD. After adjusting for smoking status and alcohol intake, serum folate concentration modeled continuously was not significantly associated with any epigenetic aging biomarker (*p* > 0.05); however, the third vs. first tertile of serum folate was positively associated with DNAmTL EAD (B (95% CI) = 0.04 kb (0.01, 0.07); *p* = 0.043) (**Supplemental Table 4**). After adjusting for cystatin C, serum folate was associated with GrimAge2 EAD (B (95% CI) = −0.46 years per doubling (−0.91, −0.01); *p* = 0.047), but not DunedinPoAm or DNAmTL EAD. In analyses stratified by smoking status, serum folate concentration was negatively associated with GrimAge2 EAD (B (95% CI) = −1.42 years per doubling (−2.50, −0.34); *p* = 0.014) and DunedinPoAm (B (95% CI) = −0.24 SDs (−0.45, −0.03); *p* = 0.031) among current smokers (*i.e.*, the group with significantly lower serum folate concentration), but not among never or former smokers, although the serum folate and smoking interaction term was not significant (**Supplemental Table 5**).

Before controlling for smoking status and alcohol intake, RBC folate concentration was positively associated with PhenoAge EAD (*B* = 0.88 years per doubling; 95% CI = −0.10, 1.85; *p* = 0.07), although this association did not reach statistical significance (Figure 2 and **Supplementary Table 3)**. With adjustment for smoking and alcohol, the association of RBC folate concentration and PhenoAge EAD increased (*B* = 1.06 years per doubling; 95% CI = −0.03, 2.14; *p* = 0.06), and there was a trend toward a positive association with GrimAge2 EAD (*B* = 0.59 years per doubling; 95% CI = −0.08, 1.26; *p* = 0.08), and DunedinPoAm (*B* = 0.11 SDs per doubling; 95% CI = −0.08, 1.26; *p* = 0.08). RBC folate concentration among former smokers were not significantly different from those of never smokers. However, among former smokers, RBC folate concentration was significantly associated with GrimAge2 EAD (*B* = 1.15 years per doubling; 95% CI = 0.40, 1.89; *p* = 0.006) and DunedinPoAm (*B* = 0.15 SDs per doubling; 95% CI = 0.00, 0.30; *p* = 0.047) (**Supplemental Table 5**). Associations were not significant among never or current smokers; however, the interaction term was not significant.

B12 concentration and tertiles were not significantly associated with any epigenetic aging biomarker (*p* > 0.05).

### Associations of homocysteine and methylmalonic acid with epigenetic aging biomarkers

Associations of markers of OCM deficiencies (*i.e.*, Hcy and MMA) with epigenetic aging biomarkers are shown in Figure 3 and **Supplementary Table 3**. Hcy concentration was positively associated with PhenoAge EAD (*B* (95% CI) = 1.05 years per doubling (0.06, 2.04); *p* = 0.039), GrimAge2 EAD (*B* (95% CI) = 1.93 years per doubling (1.16, 2.71); *p* < 0.001), and DunedinPoAm (*B* (95% CI) = 0.26 SDs per doubling (0.10, 0.41); *p* = 0.003). After adjusting for smoking status and alcohol intake, associations of Hcy with GrimAge2 EAD and DunedinPoAm remained significant but attenuated (GrimAge2 EAD: *B* (95% CI) = 1.32 years per doubling (0.69, 1.94); *p* = 0.001 and DunedinPoAm: *B* (95% CI) = 0.14 SDs per doubling (0.00, 0.27); *p* = 0.048). With cystatin C adjustment, the association with GrimAge2 EAD was further attenuated (*B* (95% CI) = 0.82 years per doubling (0.05, 1.60); *p* = 0.040), and the association with DunedinPoAm was null. Hyperhomocysteinemia vs. normal Hcy range was associated with greater PhenoAge EAD (*B* (95% CI) = 1.97 years (0.54, 3.40); *p* = 0.012) and GrimAge2 EAD (*B* (95% CI) = 2.00 years (0.83, 3.17); *p* = 0.003) (**Supplemental Table 4**). In stratified analyses, Hcy concentration was significantly and positively associated with GrimAge2 EAD among former smokers (*B* (95% CI) = 1.47 years year doubling (0.47, 2.48); *p* = 0.008) and current smokers (*B* (95% CI) = 1.76 years per doubling (0.50, 3.03); *p* = 0.010), but not among never smokers (*B* (95% CI) = 0.82 years per doubling (−0.11, 1.75); *p* = 0.08), with significant interaction between Hcy concentration and former vs. never smoking (*p_int_* = 0.027) (Figure 4 and **Supplemental Table 5**).

**Figure 3:**
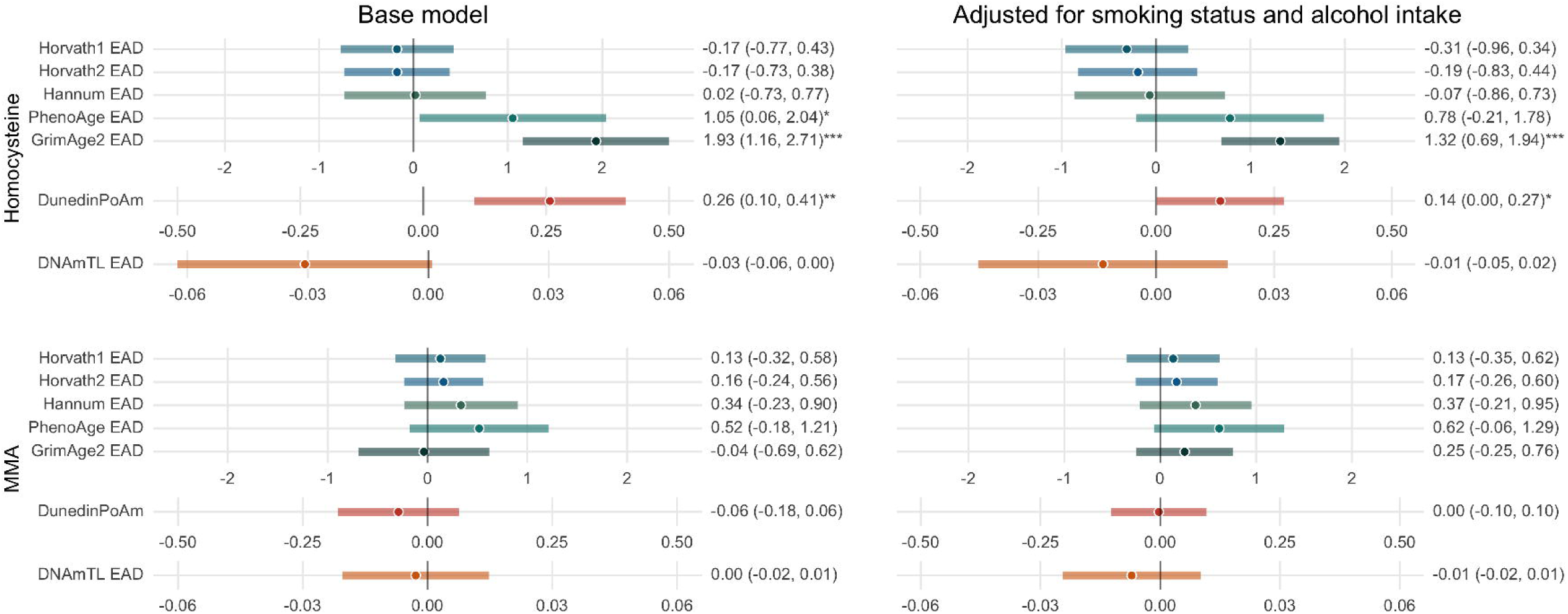
Associations of markers of one-carbon metabolism (OCM) deficiencies with epigenetic aging biomarkers. Effect estimates (95% confidence intervals (CIs)) and *p*-values are shown for a doubling in concentration of each OCM-related compound. Results are from weighted generalized linear regression models adjusted for age, age^2^, sex, race and ethnicity, BMI, education level, occupation, and poverty to income ratio. Effect estimates for Horvath1, Horvath2, Hannum, PhenoAge, and GrimAge2 EAD are in years; effect estimates for DunedinPoAm are in standard deviations; and effect estimates for DNAmTL EAD are in kilobases. EAD = epigenetic age deviation; MMA = methylmalonic acid. * *p* < 0.05; ** *p* < 0.01; *** *p* < 0.001.

**Figure 4:**
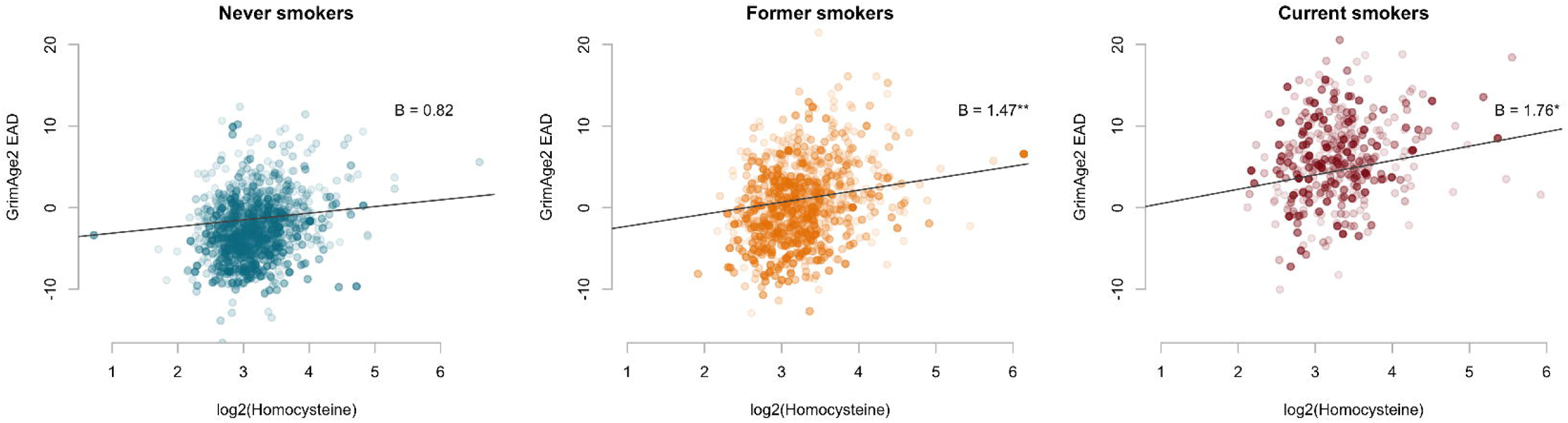
Scatter plots and associations of homocysteine concentration with GrimAge2 epigenetic age deviation (EAD). The transparency of points corresponds to their survey weights. Estimates are from weighted generalized linear regression models and shown for average age, BMI, alcohol intake, and poverty to income ratio, and for the reference level of race and ethnicity, education level, and occupation. Interaction p-value for never smokers vs. former smokers = 0.27. * *p* < 0.05; ** *p* < 0.01.

MMA concentration was not associated with the epigenetic aging biomarkers in our primary analyses. However, among former smokers, MMA was positively associated with PhenoAge EAD (*B* (95% CI) = 0.88 years per doubling (0.16, 1.60); *p* = 0.021).

### Associations with GrimAge2 components

GrimAge2 is calculated as a weighted linear combination of DNA methylation-based surrogates for 9 plasma proteins and smoking pack years. Therefore, to evaluate if individual components of GrimAge2 were driving the associations of serum folate, RBC folate, and Hcy with GrimAge2 EAD, we evaluated associations of each of the components separately. For serum folate concentration, only the association with the surrogate for smoking pack years was significant, even after adjusting for self-reported smoking status (Z-score scale) (*B* (95% CI) = −0.07 per doubling (−0.13, −0.01); *p* = 0.033) (**Table 2**). RBC folate concentration was significantly associated with GDF15 (*B* (95% CI) = 0.07 per doubling (0.01, 0.03); *p* = 0.023), PAI1 (*B* (95% CI) = 0.14 per doubling (0.01, 0.27); *p* = 0.038), and log(CRP) (*B* (95% CI) = 0.23 per doubling (0.11, 0.35); *p* = 0.002). Hcy concentration was positively associated with five GrimAge2 components: B2M (*B* (95% CI) = 0.14 per doubling (0.06, 0.22); *p* = 0.002), Cystatin C (*B* (95% CI) = 0.09 per doubling (0.02, 0.16); *p* = 0.021), TIMP1 (*B* (95% CI) = 0.08 per doubling (0.03, 0.13); *p* = 0.005), ADM (*B* (95% CI) = 0.17 per doubling (0.06, 0.29); *p* = 0.007), and smoking pack years (*B* (95% CI) = 0.15 per doubling (0.02, 0.28); *p* = 0.030).

**Table 2:**
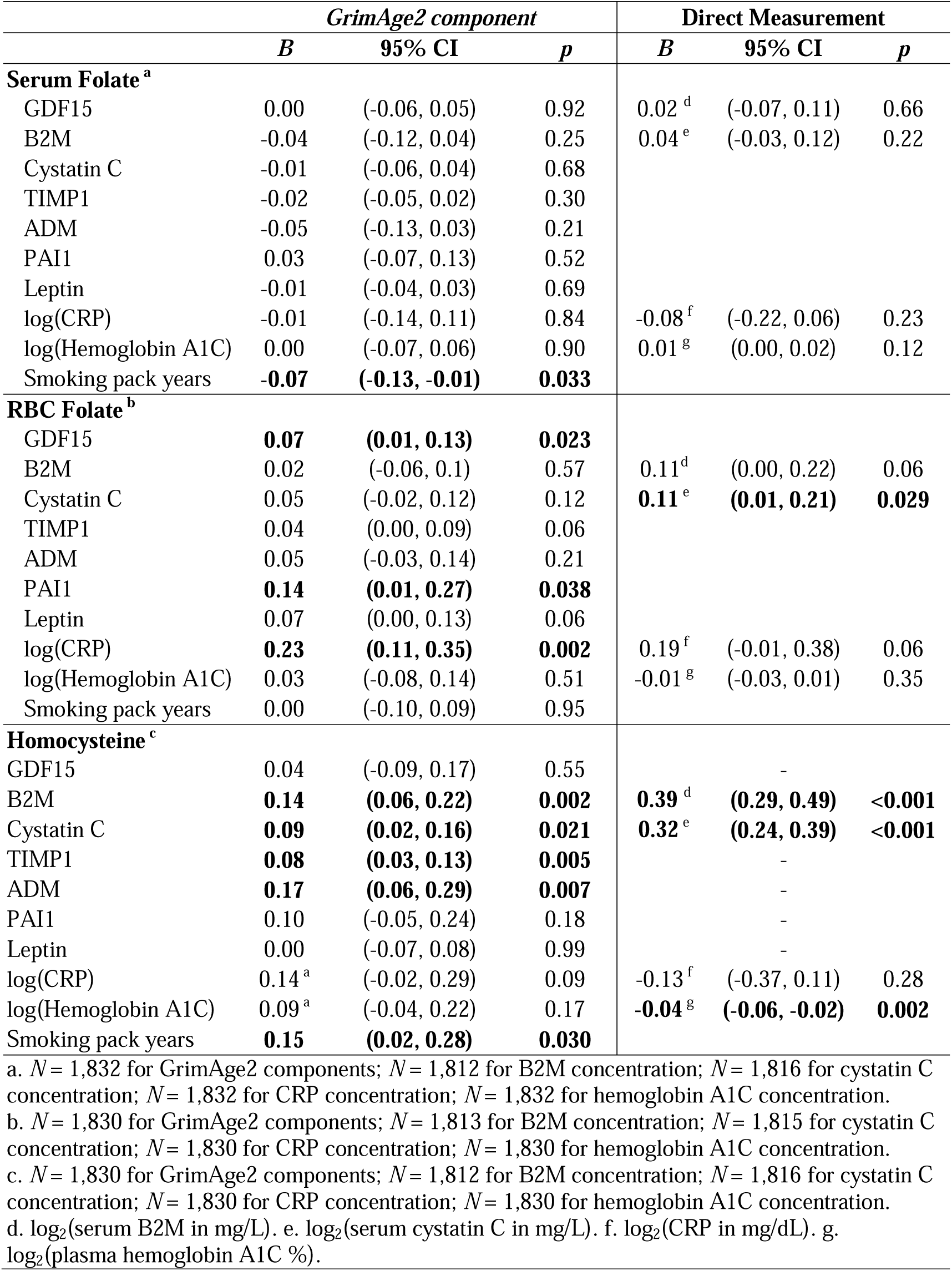
Associations of serum folate, red blood cell (RBC) folate, and homocysteine with GrimAge2 components and measured proteins. Effect estimates (95% confidence intervals (CIs)) and *p*-values are shown for a doubling in homocysteine concentration. GrimAge2 components are expressed on a Z-score scale. Results are from weighted generalized linear regression models adjusted for age, age^2^, sex, race and ethnicity, BMI, smoking status, alcohol intake, education level, occupation, and poverty to income ratio

Laboratory measures for serum B2M and cystatin C concentrations, CRP concentration, and hemoglobin A1C were also available in these NHANES cycles. Within our sample, all GrimAge2 components were moderately correlated with their measured counterparts (without survey weighting: B2M: *r* = 0.25; cystatin C: *r* = 0.22; CRP: *r* = 0.28; hemoglobin A1C: *r* = 0.50; *p* < 0.001). Therefore, we also tested if Hcy concentration was significantly associated with measured values (**Table 2**). Overall, results were consistent with those of the GrimAge2 components. However, RBC folate concentration was positively associated log_2_(cystatin C) (*B* (95% CI) = 0.11 per doubling (0.01, 0.21); *p* = 0.029) Hcy concentration was also associated with lower log_2_(hemoglobin A1C) (*B* (95% CI) = −0.04 per doubling (0.06, 0.02)).

### Sensitivity analyses

In sensitivity analyses adjusting for estimated cell type proportions, results were similar to those of our primary analyses but attenuated (**Supplemental Table 6**). The association of Hcy concentration with GrimAge2 EAD remained significant (*B* (95% CI) = 1.13 years per doubling (0.52, 1.75); *p* = 0.007). Analyses using imputed covariate data were also largely consistent with our primary analyses (**Supplemental Table 7**). A doubling in Hcy concentration was associated with 1-year greater PhenoAge EAD (95% CI = 0.13, 1.87, *p* = 0.029), 1.44 years greater GrimAge2 EAD (95% CI = 0.87, 2.00, *p* <0.001), and 0.17 SD greater DunedinPoAm (95% CI = 0.04, 0.31, *p* = 0.019).

## Discussion

We tested associations of OCM-related biomarkers with epigenetic aging biomarkers in NHANES, a nationally representative sample of adults in the US where DNA methylation was assayed on the population age 50 and older. The strongest and most robust associations were with EAD measured by second generation clocks. We provide evidence that greater concentration of Hcy, an indicator of OCM deficiencies, is associated with greater GrimAge2 EAD and DunedinPoAm. For each doubling of Hcy concentration, GrimAge2 EAA was 1.32 years greater and DunedinPoAm was 0.14 SDs greater. This association persisted after adjustment for smoking status and alcohol intake – folate antagonists and behaviors associated with poor diet quality. We also found that serum folate concentration was significantly associated with lower GrimAge2 EAD and DunedinPoAm and greater DNAmTL EAD; however, associations were imprecise after adjusting for smoking status and alcohol intake. Similarly, the association of Hcy concentration with greater PhenoAge EAD was attenuated after controlling for smoking and alcohol. It is known that smoking and alcohol intake are associated with increased EAD,^69–71^ and changes in OCM-related biomarkers may be one mechanism through which this occurs.

While our study did not directly investigate clinical endpoints, our findings support previous research linking elevated Hcy to adverse age-related health outcomes, including cognitive impairment and decline,^18^ cardiovascular disease incidence and^17^ mortality^17^ and all-cause mortality.^17^ Particularly relevant to the current study, in NHANES 1999–2006, Hcy has been associated with increased all-cause mortality and CVD mortality.^72^ GrimAge2 is an epigenetic clock developed to estimate mortality risk.^26^ GrimAge2 is calculated as the weighted linear combination of sex, age, and DNA methylation surrogates for smoking pack years and 9 plasma proteins (adrenomedullin (ADM), B2M, cystatin C, growth differentiation factor 15 (GDF-15), leptin, CRP, hemoglobin A1C, plasminogen activation inhibitor 1 (PAI-1), and tissue inhibitor metalloproteinase 1 (TIMP-1)). In addition to the overall positive association of Hcy concentration and hyperhomocysteinemia with GrimAge2 EAD, we found that Hcy was associated with greater DNA methylation surrogates of B2M (a biomarker of cardiovascular disease, kidney function, and inflammation^73^), cystatin C (a biomarker of kidney function^74^), TIMP1 (a regulator of cell growth^75^ and inflammation^76^), ADM (a vasodilator and prognostic marker in CVD^77^), and smoking pack years. In addition, analyses of directly measured protein levels were consistent with our findings on epigenetic estimates. In stratified analyses, we found evidence that Hcy has a stronger association with GrimAge2 EAD among former and current smokers compared to never smokers.

DunedinPoAm is an epigenetic biomarker that estimates the rate of change of 18 biomarkers capturing organ-system integrity and was trained on data over a 12-year span.^28^ The biomarkers included both blood biochemistries (*e.g.*, hemoglobin A1C, CRP, lipoprotein (a), HDL cholesterol) and functional and cardiometabolic measurements (*e.g.*, BMI, cardiorespiratory fitness, pulmonary function). Greater DunedinPoAm has been associated with greater physical and cognitive decline, morbidity, and mortality.^28^ In our study, Hcy was positively associated with DunedinPoAm and was robust to smoking and alcohol adjustment. However, this association was null after adjustment for cystatin C, suggesting that kidney function may be involved in the mechanism linking Hcy to DunedinPoAm.

We also found significant negative associations of serum folate with GrimAge2 EAD and DunedinPoAm before adjusting for smoking and alcohol. After smoking and alcohol intake adjustment, associations remained negative but did not reach statistical significance. Stratified analyses suggested that folate may be protective against increased DunedinPoAm and GrimAge2 EAD among smokers, although we did not find significant Hcy and smoking interaction. However, our analyses of smoking-specific effects are limited by small sample size (*N* = 811 never smokers, *N* = 732 former smokers, and *N* = 287 current smokers with complete serum folate and complete covariate data).

In contrast to our hypothesis that higher folate concentration would be protective against epigenetic aging, we observed a trend toward a positive association between RBC folate concentration and epigenetic aging measured by second generation clocks and DunedinPoAm. Although these associations did not reach statistical significance in the NHANES sample overall, associations of RBC folate with GrimAge2 EAD and DunedinPoAm were positive and significant among former smokers. Serum folate and RBC folate are both indicators of folate deficiency and were significantly correlated in our study. Although RBC folate is reflective of long-term folate status, measurement of RBC folate is more costly, has greater analytical variability, and has not shown diagnostic value in clinical settings beyond that of serum folate concentrations.^78^ We hypothesize that RBC folate concentration may also capture pathophysiological changes beyond serum folate measurements. Red blood cell abnormalities, including megaloblastic anemia, macrocytosis, and increased mean cell volume and red cell distribution width, are associated with folate and B12 deficiencies.^79^ However, cell distribution width has also been positively associated with inflammatory markers and oxidative stress,^80,81^ and RBCs may be involved in inflammatory processes due to their cytokine binding capacity.^82^ The second generation clocks and DunedinPoAm are trained on biomarkers of inflammation, such as CRP (PhenoAge, GrimAge2, DunedinPoAm), white blood cell count (DunedinPoAm), mean cell volume (PhenoAge), and red blood cell distribution width (PhenoAge), and therefore these clocks may be sensitive to inflammatory-related variations in RBCs. Furthermore, RBC folate concentration was positively associated with cystatin C, both in our study and a previous analysis of NHANES.^58^ The relationship between RBC folate and the second-generation clocks may also be reflecting impaired renal function related in aging and chronic disease development. These explanations are supported by our analyses of RBC folate concentration with the GrimAge2 components and measured proteins related to inflammation and renal function.

Associations of plasma folate, B12, B6, and Hcy with Horvath1 EAD, PhenoAge EAD, and GrimAge EAD have previously been investigated in the Veterans Affairs Normative Aging Study, a cohort of older community-dwelling men (*N* = 715).^83^ This study used Bayesian Kernel Machine Regression (BKMR), a mixture approach that allows for the selection of statistically important independent variables. B6 was identified as important for predicting PhenoAge EAD with a negative direction of association, and folate was identified as important for predicting GrimAge EAD and PhenoAge EAD with positive directions of associations. Our findings were not consistent with these results, possibly due to differences in populations studied and analytical differences. BKMR produces single-exposure effect estimates holding all other exposures at a fixed percentile. Given the complex relationship between nutritional factors and OCM metabolites, including an antagonistic relationship between one-carbon donors and Hcy, it may be difficult to interpret effects of folate when B12, B6, and Hcy are evaluated at a common percentile.

Associations of OCM-related compounds and epigenetic aging have also been analyzed in a supplementation study among older adults with mild cognitive impairment (*N* = 217). This study reported a positive correlation between baseline plasma Hcy and rate of aging (*i.e.*, epigenetic age divided by chronological age) measured by Horvath2, Hannum, Zhang,^84^ DunedinPACE,^85^ and Index, a commercially available epigenetic clock (Elysium Health, New York, NY, US).^86^ Furthermore, this study found evidence that supplementation with a B-vitamin complex (B6, B12, and folic acid) may decrease epigenetic age among individuals with hyperhomocysteinemia.

Understanding the relationship between OCM-related compounds and epigenetic aging biomarkers may help advance the study of nutrition and aging. Randomized controlled trials have demonstrated that folic acid supplementation lowers homocysteine levels,^87,88^ particularly among individuals with low baseline folate.^89^ However, there is less evidence for the downstream health benefits of supplementation with folic acid or other B vitamins. In a 7-year study of women with high risk of CVD (*N* = 5,442), supplementation with folic acid, B6, and B12 did not significantly reduce the risk of cardiovascular events compared to placebo, despite a significant reduction in homocysteine levels in the treatment group.^90^ Similarly, a 3-year study of adults with cerebral infarction (*N* = 3,680) found no association of supplementation with folic acid, B6, and B12 with risk of stroke, although lower Hcy was associated with reduced risk of coronary heart disease events.^91^ Meta-analyses of folic acid trials concluded that supplementation may reduce the risk of stroke but not mortality^92^, and that reductions in CVD were greatest among individuals with low baseline folate and high reductions in Hcy.^93^ Inconsistent results have been found for studies of folic acid and B vitamin supplementation and cognitive function;^94,95^ effects may also be modified by baseline nutritional status.^96^ Variation in results of clinical trials may be due in part to differences in outcomes measured and follow-up time. Although these studies focused on clinical outcomes, future research may leverage epigenetic aging biomarkers as surrogate endpoints in clinical trials that represent intermediate changes in complex biological pathways.^97^ Our results support the use of some of these biomarkers to test and monitor intervention prior to disease diagnosis or onset.

Our study was strengthened by its large sample size and population representative of middle-aged and older adults in the US. We were able to analyze associations with multiple OCM-related compounds and epigenetic clocks to better understand nuances in the relationships between OCM and aging-related biomarkers. However, we were limited by cross-sectional data and the inability to evaluate the effects of changes in OCM status on long-term epigenetic aging and health. A portion of our sample had missing data on covariates and therefore our primary analyses were restricted to complete cases; however, sensitivity analyses using imputed data were consistent. Additionally, we were limited by the nutritional biomarker data available for NHANES. A more comprehensive study including additional OCM-related micronutrients (*e.g.*, choline, betaine and B6) may provide further insights to the relationship between nutrition and epigenetic aging. Our study was also unable to analyze effect modification by genetic factors that affect OCM, such as variants in methylenetetrahydrofolate reductase (*MTHFR*), the enzyme that processes folate. Multiple testing was not conducted, and therefore some findings might be due to chance. However, we focused on precision of estimates along with pre-hypothesized associations, making such adjustments non-essential. Furthermore, our study was limited to a sample representative of the US adult population, and therefore findings may not be generalizable to populations with different dietary habits, nutritional status, lifestyle habits, and ages.

## Conclusion

As the application of epigenic aging biomarkers becomes more common in research and clinical settings, it is important to understand how intervenable factors, such as nutrition, influence epigenetic aging. In this study of a nationally representative sample of middle-aged and older adults, we found that Hcy concentration is positively associated with epigenetic aging measured by GrimAge2 and DunedinPoAm, particularly among former and current smokers. We also provide evidence that serum folate is associated with lower GrimAge2 EAD and DunedinPoAm and higher DNAmTL EAD. These associations were attenuated after adjusting for smoking status and alcohol intake, suggesting that changes in OCM-related biomarkers may be one mechanism through behavioral factors are related to EAD. Future studies should address the long-term health consequences of OCM-related epigenetic aging and investigate associations in subpopulations such as smokers that may have greater benefit from nutritional interventions.

## Data Availability

All datasets analyzed in the current study are publicly available from the NHANES website.

https://wwwn.cdc.gov/nchs/nhanes/

## Declaration of generative AI in scientific writing

No writing AI assistance was utilized in the writing or production of this manuscript.

## Data sharing

All datasets analyzed in the current study are publicly available from the NHANES website.

## Abbreviations

ADM: adrenomedullin
B2M: β-2 microglobulin
BKMR: Bayesian Kernel Machine Regression
BMI: body mass index
BMIQ: beta mixture quantile
CI: confidence interval
CpG: cytosine-guanine dinucleotide
CRP: C-reactive protein
CVD: cardiovascular disease
DNAmTL: DNA methylation telomere length
DunedinPoAm: Dunedin Pace-of-Aging
EAD: epigenetic age deviation
Hcy: homocysteine
HHS: Department of Health and Human Services
GDF-15: growth differentiation factor 15
MAE: median absolute error
MMA: methylmalonic acid
NCHS: National Center for Health Statistics
NHANES: National Health and Nutrition Examination Survey
OCM: one-carbon metabolism
PAI-1: plasminogen activation inhibitor 1
PIR: poverty to family income ratio
QC: quality control
RBC: red blood cell
SAH: *S*-adenosylhomocysteine
SAM: *S*-adenosylmethionine
SD: standard deviation
TIMP-1: tissue inhibitor metalloproteinase 1
US: United States

