## Supplemental Material for "One-carbon metabolism-related compounds are associated with epigenetic aging biomarkers: Results from National Health and Nutrition Examination Survey (NHANES) 1999-2002"

**Supplemental Figure 1: Participant flowchart.**

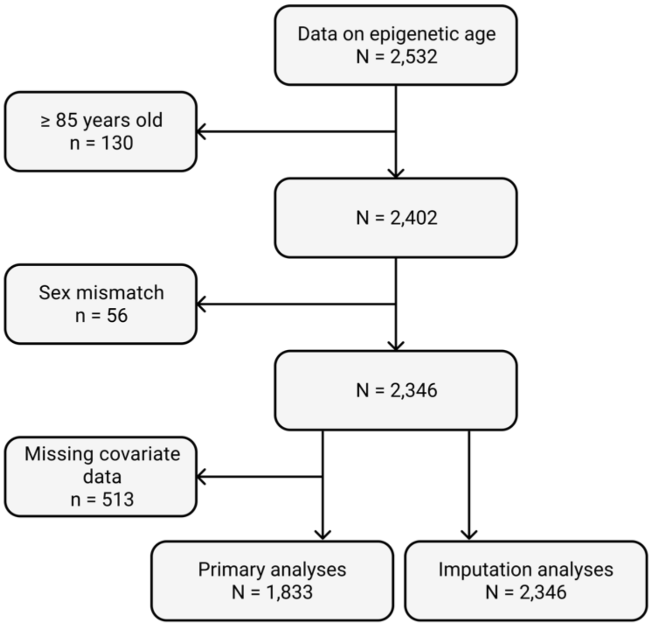

**Supplemental Figure 2: Performance of epigenetic aging biomarkers.** The identity line is plotted in orange for Horvath1, Horvath2, Hannum, PhenoAge, and GrimAge2 clocks. MAE = median absolute error.

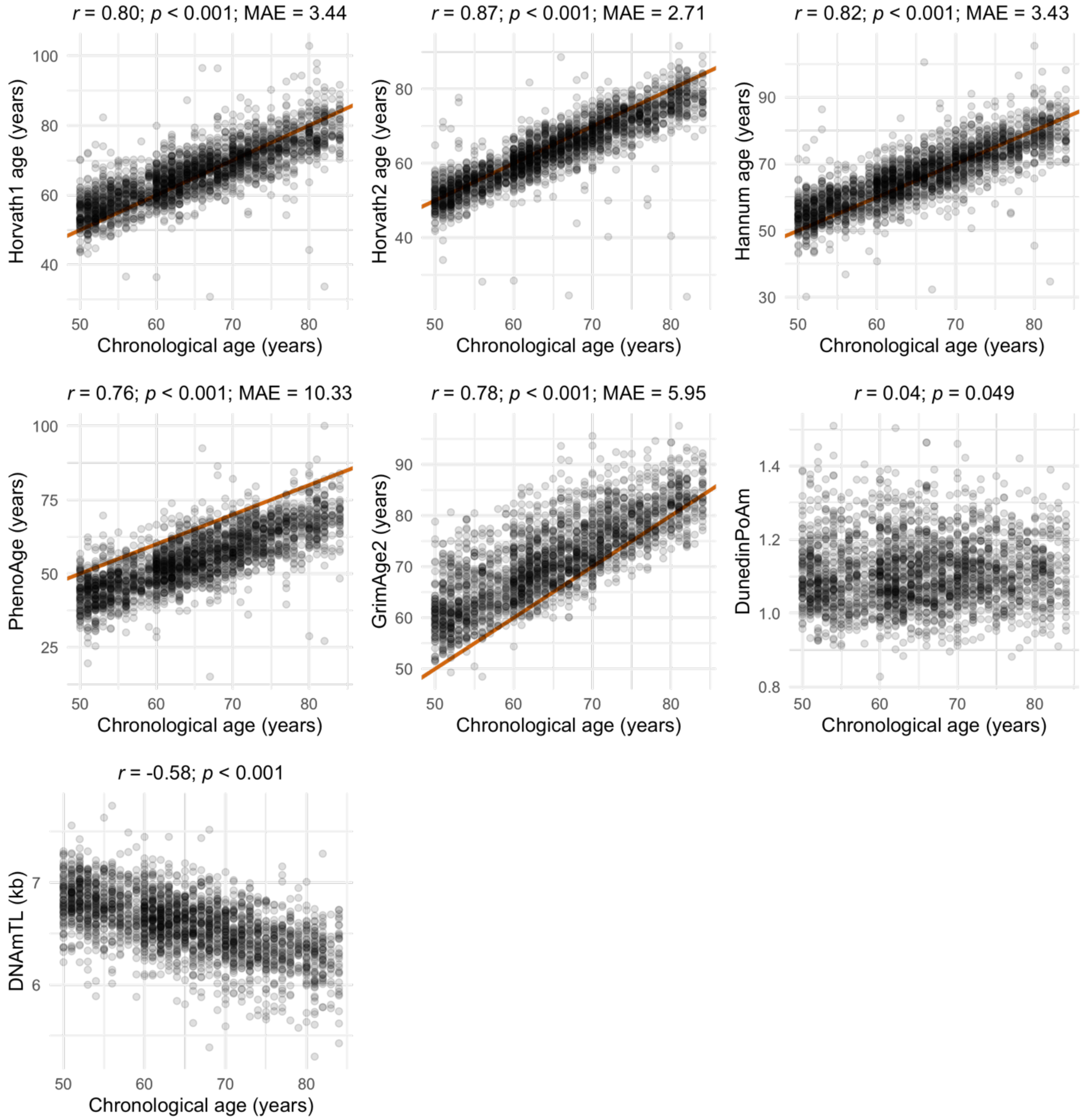

**Supplemental Figure 3:** Correlations of one-carbon metabolism-related compounds. MMA = methylmalonic acid; RBC = red blood cell.

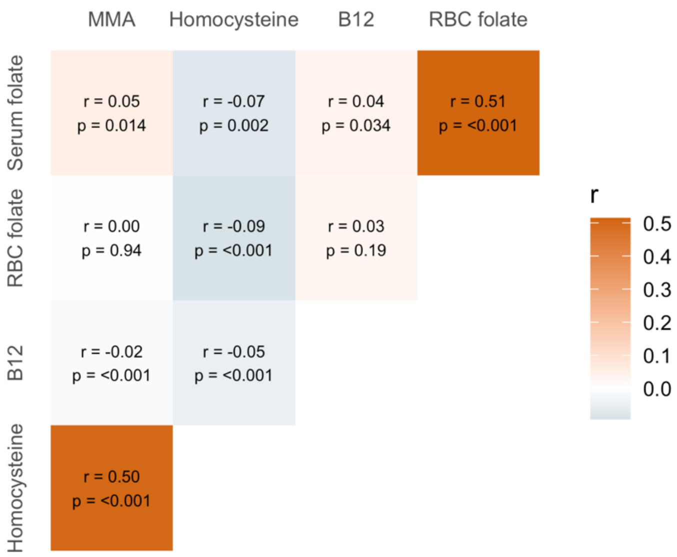

**Supplemental Table 1: Participant characteristics stratified by self-reported smoking status.** Only participants with complete covariate data are shown. P-values are from Wilcoxon rank sum tests or chi-squared tests comparing former or current smokers to never smokers.

| | **Never smokers** | **Former smokers** | | **Current smokers** | |
|---|---|---|---|---|---|
| | **n (%) or mean (SD)** | **n (%) or mean (SD)** | ***p*** | **n (%) or mean (SD)** | ***p*** |
| | 812 | 734 | | 287 | |
| Male, n (%) | 320 (39.4%) | 490 (66.8%) | <0.001 | 173 (60.3%) | <0.001 |
| Age at screening, mean (SD) | 65.4 (9.4) | 65.8 (9.2) | 0.42 | 61.4 (8.2) | <0.001 |
| Race and ethnicity, n (%) | | | 0.03 | | 0.16 |
| Mexican American | 233 (28.7%) | 211 (28.7%) | | 68 (23.7%) | |
| Other Hispanic | 48 (5.9%) | 35 (4.8%) | | 22 (7.7%) | |
| Non-Hispanic White | 322 (39.7%) | 341 (46.5%) | | 106 (36.9%) | |
| Non-Hispanic Black | 178 (21.9%) | 129 (17.6%) | | 80 (27.9%) | |
| Other race and Multiracial | 31 (3.8%) | 18 (2.5%) | | 11 (3.8%) | |
| BMI (kg/m2), mean (SD) | 29.2 (5.96) | 29.1 (5.68) | 0.95 | 27.3 (6.17) | <0.001 |
| Alcohol intake, drinks/day, mean (SD) | 0.2 (0.6) | 0.4 (0.9) | <0.001 | 0.8 (1.6) | <0.001 |
| Serum folate (ng/mL), mean (SD) | 17.4 (11.9) | 17.1 (14.4) | 0.11 | 14.0 (8.9) | <0.001 |
| Missing, n (%) | 1 (0.1%) | 0 (0%) | | 0 (0%) | |
| Folate deficiency [a] | 0 (0%) | 1 (0.1%) | 0.96 | 2 (0.7%) | 0.12 |
| Red blood cell folate (ng/mL RBC), mean (SD) | 341 (166) | 335 (164) | 0.39 | 290 (139) | <0.001 |
| Missing, n (%) | 1 (0.1%) | 2 (0.3%) | | | |
| Vitamin B12 (pg/mL), mean (SD) | 655 (1,900) | 990 (8,410) | 0.001 | 540 (524) | 0.004 |
| Missing, n (%) | 1 (0.1%) | 0 (0%) | | 0 (0%) | |
| Vitamin B12 insufficiency, n (%) [b] | 112 (13.8%) | 111 (15.1%) | 0.51 | 46 (16.0%) | 0.41 |
| Vitamin B12 deficiency, n (%) [c] | 9 (1.1%) | 6 (0.8%) | 0.74 | 3 (1.0%) | 1.00 |
| Homocysteine (umol/L), mean (SD) | 9.5 (4.8) | 10.2 (4.7) | <0.001 | 10.8 (6.0) | <0.001 |
| Missing, n (%) | 1 (0.1%) | 2 (0.3%) | | 0 (0%) | |
| Hyperhomocysteinemia [d] | 53 (6.5%) | 60 (8.2%) | 0.25 | 30 (10.5%) | 0.043 |
| Methylmalonic acid (umol/L), mean (SD) | 0.2 (0.2) | 02 (0.1) | 0.7 | 0.2 (0.2) | 0.12 |
| Missing, n (%) | 1 (0.1%) | 4 (0.5%) | | 0 (0%) | |

a. Serum folate ≤ 3 ng/mL. b. B12 < 300 pg/mL. c. < 150 pg/mL. d. > 15 umol/L.

**Supplemental Table 2: Associations of self-reported smoking status and alcohol intake with OCM-related compounds.** Effect estimates (95% confidence intervals (CIs)) and *p*-values are shown for self-reported former and current smoking vs. never smoking. Results are from weighted generalized linear regression models for log2-transformed concentrations adjusted for age, sex, race and ethnicity, BMI, education level, occupation, and poverty to income ratio. Alcohol intake is calculated as the average number of drinks/day during the past 12 months.

| | **Exposure** | ***B*** | **95% CI** | ***p*** | ***N*** |
|---|---|---|---|---|---|
| Serum folate | Smoking status | | | | 1,894 |
| | Former vs. never | -0.02 | (-0.13, 0.08) | 0.63 | |
| | Current vs. never | -0.27 | (-0.41, -0.13) | 0.001 | |
| | Alcohol intake | -0.04 | (-0.08, 0.00) | 0.06 | 1,832 |
| RBC folate | Smoking status | | | | 1,891 |
| | Former vs. never | -0.03 | (-0.11, 0.04) | 0.39 | |
| | Current vs. never | -0.24 | (-0.35, -0.14) | <0.001 | |
| | Alcohol intake | 0.001 | (-0.01, 0.03) | 0.45 | 1,830 |
| B12 | Smoking status | | | | 1,894 |
| | Former vs. never | 0.03 | (-0.09, 0.15) | 0.61 | |
| | Current vs. never | -0.10 | (-0.23, 0.03) | 0.11 | |
| | Alcohol intake | -0.06 | (-0.11, -0.01) | 0.016 | 1.832 |
| Hcy | Smoking status | | | | 1,892 |
| | Former vs. never | 0.01 | (-0.07, 0.09) | 0.75 | |
| | Current vs. never | 0.13 | (0.05, 0.21) | 0.003 | |
| | Alcohol intake | 0.07 | (0.02, 0.11) | 0.007 | 1,830 |
| MMA | Smoking status | | | | 1,890 |
| | Former vs. never | 0.00 | (-0.11, 0.11) | 0.96 | |
| | Current vs. never | -0.16 | (-0.27, -0.05) | 0.008 | |
| | Alcohol intake | 0.00 | (-0.04, 0.04) | 0.98 | 1,828 |

RBC = red blood cell; Hcy = homocysteine; MMA = methylmalonic acid.

**Supplemental Table 3: Associations of OCM-related compounds with epigenetic aging biomarkers.** Effect estimates (95% confidence intervals (CIs)) and *p*-values are shown for a doubling in concentration of each OCM-related compound. Results are from weighted generalized linear regression models adjusted for age, age$^2$, sex, race and ethnicity, BMI, education level, occupation, and poverty to income ratio.

| | | **Not adjusted for smoking status** | | | | **Adjusted for smoking status and alcohol intake** | | | | **Adjusted for smoking status, alcohol intake, and cystatin C** | | | |
|---|---|---|---|---|---|---|---|---|---|---|---|---|---|
| | | ***B*** | **95% CI** | ***p*** | ***N*** | ***B*** | **95% CI** | ***p*** | ***N*** | ***B*** | **95% CI** | ***p*** | ***N*** |
| Serum folate | Horvath1 EAD (yrs) | 0.16 | (-0.26, 0.59) | 0.43 | 1,897 | 0.23 | (-0.22, 0.67) | 0.29 | 1,832 | 0.16 | (-0.28, 0.59) | 0.43 | 1,816 |
| | Horvath2 EAD (yrs) | -0.11 | (-0.44, 0.21) | 0.47 | | -0.11 | (-0.44, 0.22) | 0.48 | | -0.19 | (-0.50, 0.12) | 0.20 | |
| | Hannum EAD (yrs) | 0.22 | (-0.25, 0.69) | 0.32 | | 0.29 | (-0.18, 0.77) | 0.20 | | 0.19 | (-0.22, 0.6) | 0.33 | |
| | PhenoAge EAD (yrs) | -0.14 | (-0.84, 0.55) | 0.66 | | 0.00 | (-0.74, 0.74) | 1.00 | | -0.14 | (-0.74, 0.46) | 0.61 | |
| | GrimAge2 EAD (yrs) | **-0.82** | **(-1.40, -0.23)** | **0.010** | | -0.36 | (-0.92, 0.19) | 0.18 | | **-0.46** | **(-0.91, -0.01)** | **0.047** | |
| | DunedinPoAm (SDs) | **-0.13** | **(-0.22, -0.03)** | **0.015** | | -0.04 | (-0.14, 0.05) | 0.36 | | -0.05 | (-0.14, 0.04) | 0.21 | |
| | DNAmTL EAD (kb) | **0.02** | **(0.00, 0.04)** | **0.037** | | 0.01 | (-0.01, 0.03) | 0.24 | | 0.02 | (0.00, 0.04) | 0.10 | |
| RBC folate | Horvath1 EAD (yrs) | 0.28 | (-0.45, 1.00) | 0.42 | 1,894 | 0.35 | (-0.44, 1.13) | 0.35 | 1,830 | 0.23 | (-0.59, 1.05) | 0.54 | 1,815 |
| | Horvath2 EAD (yrs) | -0.06 | (-0.73, 0.61) | 0.85 | | -0.04 | (-0.75, 0.68) | 0.91 | | -0.16 | (-0.89, 0.56) | 0.62 | |
| | Hannum EAD (yrs) | 0.53 | (-0.23, 1.29) | 0.15 | | 0.62 | (-0.18, 1.42) | 0.12 | | 0.41 | (-0.38, 1.20) | 0.27 | |
| | PhenoAge EAD (yrs) | 0.88 | (-0.10, 1.85) | 0.07 | | 1.06 | (-0.03, 2.14) | 0.06 | | 0.76 | (-0.25, 1.76) | 0.12 | |
| | GrimAge2 EAD (yrs) | -0.15 | (-0.76, 0.46) | 0.61 | | 0.59 | (-0.08, 1.26) | 0.07 | | 0.35 | (-0.24, 0.93) | 0.22 | |
| | DunedinPoAm (SDs) | -0.06 | (-0.14, 0.09) | 0.64 | | 0.11 | (-0.01, 0.22) | 0.07 | | 0.07 | (-0.04, 0.18) | 0.21 | |
| | DNAmTL EAD (kb) | 0.01 | (-0.02, 0.04) | 0.40 | | 0.00 | (-0.03, 0.03) | 0.82 | | 0.01 | (-0.02, 0.03) | 0.70 | |
| B12 | Horvath1 EAD (yrs) | 0.11 | (-0.32, 0.54) | 0.60 | 1,897 | 0.14 | (-0.31, 0.6) | 0.50 | 1,832 | 0.11 | (-0.34, 0.55) | 0.60 | 1,816 |
| | Horvath2 EAD (yrs) | 0.09 | (-0.31, 0.48) | 0.64 | | 0.09 | (-0.33, 0.51) | 0.66 | | 0.06 | (-0.35, 0.47) | 0.75 | |
| | Hannum EAD (yrs) | 0.07 | (-0.54, 0.67) | 0.82 | | 0.07 | (-0.58, 0.72) | 0.81 | | 0.02 | (-0.63, 0.67) | 0.94 | |
| | PhenoAge EAD (yrs) | -0.17 | (-0.91, 0.58) | 0.64 | | -0.16 | (-0.92, 0.61) | 0.66 | | -0.25 | (-0.98, 0.49) | 0.47 | |
| | GrimAge2 EAD (yrs) | -0.30 | (-0.72, 0.12) | 0.15 | | -0.10 | (-0.58, 0.38) | 0.64 | | -0.17 | (-0.61, 0.27) | 0.41 | |
| | DunedinPoAm (SDs) | -0.00 | (-0.11, 0.10) | 0.95 | | 0.03 | (-0.08, 0.14) | 0.57 | | 0.02 | (-0.09, 0.12) | 0.69 | |
| | DNAmTL EAD (kb) | 0.01 | (-0.01, 0.03) | 0.36 | | 0.00 | (-0.02, 0.03) | 0.68 | | 0.01 | (-0.02, 0.03) | 0.55 | |
| Hcy | Horvath1 EAD (yrs) | -0.17 | (-0.77, 0.43) | 0.54 | 1,895 | -0.31 | (-0.96, 0.34) | 0.31 | 1,830 | -0.69 | (-1.46, 0.07) | 0.07 | 1,814 |
| | Horvath2 EAD (yrs) | -0.17 | (-0.73, 0.38) | 0.51 | | -0.19 | (-0.83, 0.44) | 0.51 | | -0.60 | (-1.32, 0.12) | 0.09 | |
| | Hannum EAD (yrs) | 0.02 | (-0.73, 0.77) | 0.96 | | -0.07 | (-0.86, 0.73) | 0.85 | | -0.67 | (-1.50, 0.16) | 0.10 | |
| | PhenoAge EAD (yrs) | **1.05** | **(0.06, 2.04)** | **0.039** | | 0.78 | (-0.21, 1.78) | 0.11 | | -0.08 | (-1.29, 1.13) | 0.89 | |
| | GrimAge2 EAD (yrs) | **1.93** | **(1.16, 2.71)** | **<0.001** | | **1.32** | **(0.69, 1.94)** | **0.001** | | **0.82** | **(0.05, 1.60)** | **0.040** | |
| | DunedinPoAm (SDs) | **0.26** | **(0.10, 0.41)** | **0.003** | | **0.14** | **(0.00, 0.27)** | **0.048** | | 0.03 | (-0.12, 0.18) | 0.66 | |
| | DNAmTL EAD (kb) | -0.03 | (-0.06, 0.00) | 0.06 | | -0.01 | (-0.05, 0.02) | 0.36 | | 0.00 | (-0.04, 0.04) | 0.94 | |
| MMA | Horvath1 EAD (yrs) | 0.13 | (-0.32, 0.58) | 0.55 | 1,893 | 0.13 | (-0.35, 0.62) | 0.55 | 1,828 | -0.04 | (-0.64, 0.56) | 0.89 | 1,812 |
| | Horvath2 EAD (yrs) | 0.16 | (-0.24, 0.56) | 0.40 | | 0.17 | (-0.26, 0.6) | 0.39 | | -0.02 | (-0.49, 0.45) | 0.91 | |
| | Hannum EAD (yrs) | 0.34 | (-0.23, 0.90) | 0.22 | | 0.37 | (-0.21, 0.95) | 0.19 | | 0.09 | (-0.47, 0.65) | 0.72 | |
| | PhenoAge EAD (yrs) | 0.52 | (-0.18, 1.21) | 0.13 | | 0.62 | (-0.06, 1.29) | 0.07 | | 0.12 | (-0.58, 0.83) | 0.70 | |
| | GrimAge2 EAD (yrs) | -0.04 | (-0.69, 0.62) | 0.91 | | 0.25 | (-0.25, 0.76) | 0.29 | | -0.18 | (-0.59, 0.24) | 0.36 | |
| | DunedinPoAm (SDs) | -0.06 | (-0.18, 0.06) | 0.32 | | 0.00 | (-0.1, 0.1) | 0.95 | | -0.09 | (-0.18, 0.01) | 0.07 | |
| | DNAmTL EAD (kb) | 0.00 | (-0.02, 0.01) | 0.73 | | -0.01 | (-0.02, 0.01) | 0.38 | | 0.00 | (-0.02, 0.02) | 0.88 | |

RBC = red blood cell; Hcy = homocysteine; MMA = methylmalonic acid; EAD = epigenetic age deviation; SD = standard deviation; kb = kilobase.

**Supplemental Table 4: Associations of folate and B12 tertiles and hyperhomocysteinemia with epigenetic aging biomarkers.** Effect estimates (95% confidence intervals (CIs)) and *p*-values are shown. Results are from weighted generalized linear regression models adjusted for age, age$^2$, sex, race and ethnicity, BMI, smoking status, alcohol intake, education level, occupation, and poverty to income ratio. RBC = red blood cell.

| | | | ***B*** | **95% CI** | ***p*** | ***N*** |
|---|---|---|---|---|---|---|
| Serum folate | Tertile 2 vs. 1 | Horvath1 EAD (yrs) | -0.16 | (-1.13, 0.81) | 0.72 | 1,832 |
| | | Horvath2 EAD (yrs) | -0.34 | (-1.07, 0.39) | 0.32 | |
| | | Hannum EAD (yrs) | 0.12 | (-0.84, 1.07) | 0.79 | |
| | | PhenoAge EAD (yrs) | 0.14 | (-0.95, 1.22) | 0.78 | |
| | | GrimAge2 EAD (yrs) | -0.51 | (-1.30, 0.28) | 0.18 | |
| | | DunedinPoAm (SDs) | -0.12 | (-0.34, 0.10) | 0.24 | |
| | | DNAmTL EAD (kb) | 0.02 | (-0.01, 0.05) | 0.22 | |
| | Tertile 3 vs. 1 | Horvath1 EAD (yrs) | 0.26 | (-0.68, 1.200) | 0.55 | |
| | | Horvath2 EAD (yrs) | -0.41 | (-1.05, 0.24) | 0.19 | |
| | | Hannum EAD (yrs) | 0.26 | (-0.58, 1.1) | 0.50 | |
| | | PhenoAge EAD (yrs) | -0.25 | (-1.45, 0.94) | 0.65 | |
| | | GrimAge2 EAD (yrs) | -0.77 | (-1.57, 0.04) | 0.06 | |
| | | DunedinPoAm (SDs) | -0.09 | (-0.25, 0.08) | 0.25 | |
| | | DNAmTL EAD (kb) | **0.04** | **(0.00, 0.07)** | **0.043** | |
| RBC folate | Tertile 2 vs. 1 | Horvath1 EAD (yrs) | -0.61 | (-1.39, 0.18) | 0.11 | 1,830 |
| | | Horvath2 EAD (yrs) | -0.26 | (-0.82, 0.31) | 0.33 | |
| | | Hannum EAD (yrs) | -0.36 | (-1.07, 0.36) | 0.29 | |
| | | PhenoAge EAD (yrs) | -0.34 | (-1.55, 0.87) | 0.54 | |
| | | GrimAge2 EAD (yrs) | -0.15 | (-0.99, 0.69) | 0.70 | |
| | | DunedinPoAm (SDs) | -0.03 | (-0.23, 0.16) | 0.71 | |
| | | DNAmTL EAD (kb) | 0.02 | (-0.01, 0.06) | 0.20 | |
| | Tertile 3 vs. 1 | Horvath1 EAD (yrs) | 0.18 | (-0.75, 1.10) | 0.68 | |
| | | Horvath2 EAD (yrs) | -0.33 | (-1.01, 0.35) | 0.30 | |
| | | Hannum EAD (yrs) | 0.47 | (-0.42, 1.35) | 0.26 | |
| | | PhenoAge EAD (yrs) | 0.82 | (-0.69, 2.32) | 0.25 | |
| | | GrimAge2 EAD (yrs) | 0.31 | (-0.47, 1.10) | 0.39 | |
| | | DunedinPoAm (SDs) | 0.10 | (-0.07, 0.27) | 0.21 | |
| | | DNAmTL EAD (kb) | 0.00 | (-0.04, 0.04) | 0.93 | |
| B12 | Tertile 2 vs. 1 | Horvath1 EAD (yrs) | 0.61 | (-0.21, 1.43) | 0.13 | 1,832 |
| | | Horvath2 EAD (yrs) | 0.09 | (-0.54, 0.73) | 0.75 | |
| | | Hannum EAD (yrs) | -0.06 | (-0.86, 0.74) | 0.87 | |
| | | PhenoAge EAD (yrs) | -0.17 | (-1.23, 0.89) | 0.73 | |
| | | GrimAge2 EAD (yrs) | 0.13 | (-0.67, 0.92) | 0.73 | |
| | | DunedinPoAm (SDs) | -0.04 | (-0.23, 0.14) | 0.62 | |
| | | DNAmTL EAD (kb) | 0.01 | (-0.02, 0.04) | 0.39 | |
| | Tertile 3 vs. 1 | Horvath1 EAD (yrs) | 0.34 | (-0.46, 1.14) | 0.36 | |
| | | Horvath2 EAD (yrs) | 0.30 | (-0.45, 1.05) | 0.39 | |
| | | Hannum EAD (yrs) | 0.49 | (-0.31, 1.28) | 0.20 | |
| | | PhenoAge EAD (yrs) | -0.51 | (-1.59, 0.57) | 0.31 | |
| | | GrimAge2 EAD (yrs) | -0.18 | (-0.94, 0.58) | 0.61 | |
| | | DunedinPoAm (SDs) | 0.01 | (-0.19, 0.21) | 0.93 | |
| | | DNAmTL EAD (kb) | 0.00 | (-0.03, 0.04) | 0.79 | |

| | | | | | |
|---|---|---|---|---|---|
| Hyperhomocysteinemia | Horvath1 EAD (yrs) | -0.29 | (-1.32, 0.75) | 0.55 | 1,830 |
| | Horvath2 EAD (yrs) | 0.08 | (-0.94, 1.09) | 0.87 | |
| | Hannum EAD (yrs) | 0.04 | (-1.29, 1.36) | 0.95 | |
| | PhenoAge EAD (yrs) | **1.97** | **(0.54, 3.40)** | **0.012** | |
| | GrimAge2 EAD (yrs) | **2.00** | **(0.83, 3.17)** | **0.003** | |
| | DunedinPoAm (SDs) | 0.23 | (-0.03, 0.49) | 0.08 | |
| | DNAmTL EAD (kb) | -0.03 | (-0.10, 0.03) | 0.30 | |

RBC = red blood cell; EAD = epigenetic age deviation; SD = standard deviation; kb = kilobase.

**Supplemental Table 5: Associations of OCM-related compounds with epigenetic aging biomarkers stratified by self-reported smoking status.** Effect estimates (95% confidence intervals (CIs)) and *p*-values are shown for a doubling in concentration of each OCM-related compound. Results are from weighted generalized linear regression models adjusted for age, age$^2$, sex, race and ethnicity, BMI, alcohol intake, education level, occupation, and poverty to income ratio. Interaction *p*-values are from models of the full sample including a concentration and smoking (never smoker, former smoker, current smoker) interaction term.

| | | **Never smokers** [a] | | | **Former smokers** [b] | | | | **Current smokers** [c] | | | |
|---|---|---|---|---|---|---|---|---|---|---|---|---|
| | | ***B*** | **95% CI** | ***p*** | ***B*** | **95% CI** | ***p*** | $p_{int}$ | ***B*** | **95% CI** | ***p*** | $p_{int}$ |
| Serum folate | Horvath1 EAD (yrs) | 0.18 | (-0.4, 0.77) | 0.51 | -0.02 | (-0.78, 0.73) | 0.95 | 0.73 | 0.81 | (-0.27, 1.89) | 0.13 | 0.29 |
| | Horvath2 EAD (yrs) | -0.32 | (-0.86, 0.22) | 0.22 | -0.13 | (-0.60, 0.35) | 0.57 | 0.55 | 0.17 | (-0.89, 1.24) | 0.73 | 0.32 |
| | Hannum EAD (yrs) | 0.38 | (-0.34, 1.10) | 0.27 | 0.08 | (-0.67, 0.84) | 0.82 | 0.68 | 0.28 | (-0.90, 1.47) | 0.61 | 0.81 |
| | PhenoAge EAD (yrs) | -0.05 | (-1.08, 0.99) | 0.93 | -0.17 | (-1.13, 0.78) | 0.70 | 0.67 | 0.21 | (-1.28, 1.69) | 0.77 | 0.32 |
| | GrimAge2 EAD (yrs) | -0.04 | (-0.87, 0.79) | 0.92 | -0.24 | (-0.76, 0.28) | 0.34 | 0.84 | **-1.42** | **(-2.50, -0.34)** | **0.014** | 0.13 |
| | DunedinPoAm (SDs) | -0.03 | (-0.16, 0.10) | 0.63 | 0.01 | (-0.10, 0.13) | 0.80 | 0.45 | **-0.24** | **(-0.45, -0.03)** | **0.031** | 0.44 |
| | DNAmTL EAD (kb) | 0.01 | (-0.02, 0.04) | 0.38 | 0.01 | (-0.02, 0.05) | 0.43 | 0.66 | 0.02 | (-0.03, 0.06) | 0.44 | 0.79 |
| RBC folate | Horvath1 EAD (yrs) | 0.63 | (-0.75, 2.02) | 0.34 | -0.06 | (-0.94, 0.81) | 0.88 | 0.37 | 0.18 | (-0.86, 1.22) | 0.71 | 0.56 |
| | Horvath2 EAD (yrs) | -0.12 | (-1.38, 1.14) | 0.84 | -0.19 | (-0.84, 0.46) | 0.53 | 1.00 | 0.20 | (-1.12, 1.53) | 0.75 | 0.58 |
| | Hannum EAD (yrs) | 1.00 | (-0.53, 2.53) | 0.18 | 0.26 | (-0.55, 1.07) | 0.50 | 0.45 | -0.16 | (-1.41, 1.10) | 0.79 | 0.33 |
| | PhenoAge EAD (yrs) | 1.03 | (-0.86, 2.92) | 0.26 | 0.44 | (-0.55, 1.42) | 0.35 | 0.69 | 1.73 | (-0.41, 3.86) | 0.10 | 0.39 |
| | GrimAge2 EAD (yrs) | 0.40 | (-0.44, 1.25) | 0.32 | **1.15** | **(0.4, 1.89)** | **0.006** | 0.14 | -0.33 | (-2.16, 1.50) | 0.70 | 0.64 |
| | DunedinPoAm (SDs) | 0.11 | (-0.02, 0.23) | 0.09 | **0.15** | **(0.00, 0.30)** | **0.047** | 0.61 | -0.04 | (-0.41, 0.32) | 0.81 | 0.72 |
| | DNAmTL EAD (kb) | 0.00 | (-0.05, 0.05) | 0.92 | -0.01 | (-0.05, 0.03) | 0.62 | 0.70 | 0.03 | (-0.04, 0.1) | 0.40 | 0.46 |
| B12 | Horvath1 EAD (yrs) | 0.16 | (-0.41, 0.72) | 0.56 | 0.18 | (-0.53, 0.88) | 0.59 | 0.68 | -0.14 | (-1.51, 1.23) | 0.82 | 0.77 |
| | Horvath2 EAD (yrs) | 0.15 | (-0.43, 0.72) | 0.59 | 0.12 | (-0.46, 0.69) | 0.67 | 0.89 | -0.31 | (-1.17, 0.54) | 0.44 | 0.68 |
| | Hannum EAD (yrs) | 0.50 | (-0.22, 1.21) | 0.16 | -0.08 | (-1.14, 0.98) | 0.87 | 0.42 | -0.74 | (-1.96, 0.49) | 0.21 | 0.23 |
| | PhenoAge EAD (yrs) | 0.45 | (-0.55, 1.45) | 0.35 | -0.56 | (-1.50, 0.38) | 0.22 | 0.29 | -0.53 | (-1.80, 0.75) | 0.39 | 0.85 |
| | GrimAge2 EAD (yrs) | 0.41 | (-0.22, 1.05) | 0.18 | -0.47 | (-1.04, 0.1) | 0.10 | 0.05 | -0.41 | (-1.52, 0.70) | 0.44 | 0.39 |
| | DunedinPoAm (SDs) | 0.09 | (-0.02, 0.20) | 0.09 | 0.01 | (-0.19, 0.20) | 0.93 | 0.48 | -0.10 | (-0.30, 0.1) | 0.31 | 0.26 |
| | DNAmTL EAD (kb) | 0.02 | (-0.01, 0.04) | 0.17 | -0.01 | (-0.03, 0.02) | 0.69 | 0.06 | -0.01 | (-0.07, 0.06) | 0.77 | 0.34 |
| Hcy | Horvath1 EAD (yrs) | -0.65 | (-1.73, 0.44) | 0.22 | 0.07 | (-0.82, 0.96) | 0.86 | 0.11 | -0.25 | (-1.81, 1.31) | 0.73 | 0.41 |
| | Horvath2 EAD (yrs) | -0.44 | (-1.40, 0.52) | 0.34 | -0.13 | (-0.97, 0.72) | 0.75 | 0.37 | 0.25 | (-1.45, 1.95) | 0.75 | 0.44 |
| | Hannum EAD (yrs) | -0.54 | (-2.01, 0.93) | 0.44 | 0.10 | (-0.94, 1.15) | 0.83 | 0.30 | 0.47 | (-1.32, 2.27) | 0.58 | 0.21 |
| | PhenoAge EAD (yrs) | 0.78 | (-1.08, 2.64) | 0.38 | 0.45 | (-0.79, 1.69) | 0.44 | 0.56 | 1.15 | (-1.23, 3.52) | 0.31 | 0.37 |
| | GrimAge2 EAD (yrs) | 0.82 | (-0.11, 1.75) | 0.08 | **1.47** | **(0.47, 2.48)** | **0.008** | **0.027** | **1.76** | **(0.50, 3.03)** | **0.010** | 0.25 |
| | DunedinPoAm (SDs) | 0.08 | (-0.06, 0.22) | 0.26 | 0.19 | (-0.05, 0.42) | 0.11 | 0.21 | 0.15 | (-0.13, 0.43) | 0.27 | 0.69 |
| | DNAmTL EAD (kb) | -0.03 | (-0.08, 0.02) | 0.20 | 0.01 | (-0.03, 0.05) | 0.64 | 0.51 | -0.02 | (-0.08, 0.04) | 0.48 | 0.59 |
| MMA | Horvath1 EAD (yrs) | -0.34 | (-1.17, 0.49) | 0.40 | 0.46 | (-0.26, 1.18) | 0.19 | 0.10 | 0.57 | (-0.89, 2.03) | 0.41 | 0.29 |
| | Horvath2 EAD (yrs) | -0.10 | (-0.82, 0.61) | 0.76 | 0.10 | (-0.46, 0.66) | 0.71 | 0.37 | 1.02 | (-0.59, 2.62) | 0.19 | 0.22 |
| | Hannum EAD (yrs) | -0.18 | (-1.15, 0.78) | 0.69 | 0.58 | (-0.11, 1.26) | 0.09 | 0.16 | 1.22 | (-0.76, 3.2) | 0.20 | 0.19 |
| | PhenoAge EAD (yrs) | 0.10 | (-1.22, 1.41) | 0.87 | **0.88** | **(0.16, 1.60)** | **0.021** | 0.18 | 0.64 | (-1.79, 3.06) | 0.58 | 0.55 |
| | GrimAge2 EAD (yrs) | 0.25 | (-0.64, 1.13) | 0.56 | 0.45 | (-0.26, 1.17) | 0.19 | 0.53 | -0.38 | (-1.36, 0.60) | 0.42 | 0.25 |
| | DunedinPoAm (SDs) | -0.04 | (-0.18, 0.11) | 0.59 | 0.06 | (-0.08, 0.20) | 0.36 | 0.38 | -0.12 | (-0.38, 0.13) | 0.32 | 0.47 |
| | DNAmTL EAD (kb) | -0.01 | (-0.04, 0.03) | 0.64 | 0.00 | (-0.02, 0.02) | 0.90 | 0.97 | -0.01 | (-0.05, 0.03) | 0.50 | 0.81 |

a. *N* = 811. b. Serum folate and B12: *N* = 734; RBC folate and homocysteine: *N* = 732; MMA: *N* = 730. c. *N* = 287. RBC = red blood cell; Hcy = homocysteine; MMA = methylmalonic acid; EAD = epigenetic age deviation; SD = standard deviation; kb = kilobase.

**Supplemental Table 6: Associations of OCM-related compounds with epigenetic aging biomarkers adjusting for cell type proportions.** Effect estimates (95% confidence intervals (CIs)) and *p*-values are shown for a doubling in concentration of each OCM-related compound. Results are from weighted generalized linear regression models adjusted for age, age$^2$, sex, race and ethnicity, BMI, smoking status, alcohol intake, education level, occupation, poverty to income ratio, and estimated proportions of CB8+ T cells, CD4+ T cells, neutrophils, monocytes, B cells, and natural killer cells.

| | | ***B*** | **95% CI** | ***p*** | ***N*** |
|---|---|---|---|---|---|
| Serum folate | Horvath1 EAD (yrs) | 0.24 | (-0.33, 0.80) | 0.31 | 1,832 |
| | Horvath2 EAD (yrs) | -0.10 | (-0.50, 0.31) | 0.54 | |
| | Hannum EAD (yrs) | 0.26 | (-0.30, 0.82) | 0.27 | |
| | PhenoAge EAD (yrs) | -0.04 | (-0.79, 0.70) | 0.87 | |
| | GrimAge2 EAD (yrs) | -0.37 | (-0.93, 0.19) | 0.14 | |
| | DunedinPoAm (SDs) | -0.05 | (-0.12, 0.03) | 0.17 | |
| | DNAmTL EAD (kb) | 0.01 | (-0.02, 0.03) | 0.36 | |
| RBC folate | Horvath1 EAD (yrs) | 0.39 | (-0.61, 1.38) | 0.34 | 1,830 |
| | Horvath2 EAD (yrs) | -0.03 | (-0.93, 0.87) | 0.93 | |
| | Hannum EAD (yrs) | 0.61 | (-0.36, 1.57) | 0.15 | |
| | PhenoAge EAD (yrs) | 0.96 | (-0.31, 2.22) | 0.10 | |
| | GrimAge2 EAD (yrs) | 0.47 | (-0.25, 1.19) | 0.14 | |
| | DunedinPoAm (SDs) | 0.07 | (-0.04, 0.18) | 0.14 | |
| | DNAmTL EAD (kb) | 0.00 | (-0.04, 0.03) | 0.75 | |
| B12 | Horvath1 EAD (yrs) | -0.03 | (-0.70, 0.63) | 0.89 | 1,832 |
| | Horvath2 EAD (yrs) | -0.04 | (-0.60, 0.52) | 0.85 | |
| | Hannum EAD (yrs) | -0.16 | (-0.83, 0.50) | 0.54 | |
| | PhenoAge EAD (yrs) | -0.38 | (-1.20, 0.43) | 0.26 | |
| | GrimAge2 EAD (yrs) | -0.24 | (-0.68, 0.20) | 0.21 | |
| | DunedinPoAm (SDs) | 0.00 | (-0.08, 0.09) | 0.92 | |
| | DNAmTL EAD (kb) | 0.02 | (-0.01, 0.04) | 0.14 | |
| Hcy | Horvath1 EAD (yrs) | -0.16 | (-0.97, 0.66) | 0.62 | 1,830 |
| | Horvath2 EAD (yrs) | -0.11 | (-0.83, 0.61) | 0.69 | |
| | Hannum EAD (yrs) | 0.05 | (-0.81, 0.91) | 0.88 | |
| | PhenoAge EAD (yrs) | 0.69 | (-0.51, 1.88) | 0.18 | |
| | GrimAge2 EAD (yrs) | **1.13** | **(0.52, 1.75)** | **0.007** | |
| | DunedinPoAm (SDs) | 0.09 | (-0.03, 0.20) | 0.10 | |
| | DNAmTL EAD (kb) | -0.02 | (-0.06, 0.02) | 0.18 | |
| MMA | Horvath1 EAD (yrs) | 0.03 | (-0.54, 0.60) | 0.88 | 1,828 |
| | Horvath2 EAD (yrs) | 0.11 | (-0.36, 0.58) | 0.55 | |
| | Hannum EAD (yrs) | 0.19 | (-0.38, 0.75) | 0.42 | |
| | PhenoAge EAD (yrs) | 0.47 | (-0.23, 1.17) | 0.13 | |
| | GrimAge2 EAD (yrs) | 0.20 | (-0.34, 0.75) | 0.35 | |
| | DunedinPoAm (SDs) | -0.03 | (-0.13, 0.08) | 0.53 | |
| | DNAmTL EAD (kb) | 0.00 | (-0.02, 0.02) | 0.93 | |

RBC = red blood cell; Hcy = homocysteine; MMA = methylmalonic acid; EAD = epigenetic age deviation; SD = standard deviation; kb = kilobase.

**Supplemental Table 7: Associations of OCM-related compounds with epigenetic aging biomarkers using imputed covariate data.** Effect estimates (95% confidence intervals (CIs)) and *p*-values are shown for a doubling in concentration of each OCM-related compound. Results are from weighted generalized linear regression models adjusted for age, age$^2$, sex, race and ethnicity, BMI, smoking status, alcohol intake, education level, occupation, and poverty to income ratio. $N = 2{,}346$.

| | | ***B*** | **95% CI** | ***p*** |
|---|---|---|---|---|
| Serum folate | Horvath1 EAD (yrs) | 0.17 | (-0.21, 0.54) | 0.34 |
| | Horvath2 EAD (yrs) | -0.07 | (-0.38, 0.25) | 0.64 |
| | Hannum EAD (yrs) | 0.23 | (-0.13, 0.58) | 0.18 |
| | PhenoAge EAD (yrs) | -0.01 | (-0.51, 0.48) | 0.95 |
| | GrimAge2 EAD (yrs) | -0.34 | (-0.81, 0.12) | 0.13 |
| | DunedinPoAm (SDs) | -0.05 | (-0.14, 0.05) | 0.30 |
| | DNAmTL EAD (kb) | 0.01 | (-0.01, 0.03) | 0.18 |
| RBC folate | Horvath1 EAD (yrs) | 0.30 | (-0.45, 1.06) | 0.38 |
| | Horvath2 EAD (yrs) | 0.06 | (-0.67, 0.78) | 0.86 |
| | Hannum EAD (yrs) | 0.59 | (-0.17, 1.34) | 0.11 |
| | PhenoAge EAD (yrs) | 0.97 | (-0.03, 1.96) | 0.05 |
| | GrimAge2 EAD (yrs) | 0.54 | (-0.07, 1.15) | 0.08 |
| | DunedinPoAm (SDs) | 0.09 | (-0.03, 0.20) | 0.12 |
| | DNAmTL EAD (kb) | 0.00 | (-0.03, 0.03) | 0.93 |
| B12 | Horvath1 EAD (yrs) | 0.08 | (-0.38, 0.54) | 0.70 |
| | Horvath2 EAD (yrs) | 0.05 | (-0.40, 0.49) | 0.82 |
| | Hannum EAD (yrs) | 0.13 | (-0.52, 0.78) | 0.67 |
| | PhenoAge EAD (yrs) | -0.17 | (-0.86, 0.52) | 0.59 |
| | GrimAge2 EAD (yrs) | -0.09 | (-0.53, 0.35) | 0.64 |
| | DunedinPoAm (SDs) | 0.03 | (-0.06, 0.13) | 0.46 |
| | DNAmTL EAD (kb) | 0.01 | (-0.01, 0.03) | 0.52 |
| Hcy | Horvath1 EAD (yrs) | 0.02 | (-0.65, 0.68) | 0.96 |
| | Horvath2 EAD (yrs) | -0.06 | (-0.7, 0.58) | 0.83 |
| | Hannum EAD (yrs) | 0.18 | (-0.55, 0.91) | 0.59 |
| | PhenoAge EAD (yrs) | **1.00** | **(0.13, 1.87)** | **0.029** |
| | GrimAge2 EAD (yrs) | **1.44** | **(0.87, 2.00)** | **<0.001** |
| | DunedinPoAm (SDs) | **0.17** | **(0.04, 0.31)** | **0.019** |
| | DNAmTL EAD (kb) | -0.03 | (-0.06, 0) | 0.08 |
| MMA | Horvath1 EAD (yrs) | 0.11 | (-0.32, 0.55) | 0.56 |
| | Horvath2 EAD (yrs) | 0.15 | (-0.25, 0.55) | 0.41 |
| | Hannum EAD (yrs) | 0.27 | (-0.27, 0.81) | 0.29 |
| | PhenoAge EAD (yrs) | 0.53 | (-0.19, 1.25) | 0.13 |
| | GrimAge2 EAD (yrs) | 0.25 | (-0.17, 0.68) | 0.21 |
| | DunedinPoAm (SDs) | -0.01 | (-0.1, 0.07) | 0.73 |
| | DNAmTL EAD (kb) | -0.01 | (-0.03, 0.01) | 0.22 |

RBC = red blood cell; Hcy = homocysteine; MMA = methylmalonic acid; EAD = epigenetic age deviation; SD = standard deviation; kb = kilobase.
